# Multiomics implicate gut microbiota in altered lipid and energy metabolism in Parkinson’s disease

**DOI:** 10.1101/2021.05.29.21258035

**Authors:** Pedro A. B. Pereira, Drupad K. Trivedi, Justin Silverman, Ilhan Cem Duru, Lars Paulin, Petri Auvinen, Filip Scheperjans

## Abstract

We aimed to investigate the link between serum metabolites, gut bacterial community composition, and clinical variables in Parkinson’s disease (PD) and healthy control subjects (HC). 139 metabolite features were found to be differentially abundant between the PD and Control groups. No associations were found between metabolite features and within-PD clinical variables. The results suggest alterations in serum metabolite profiles in PD, and the results of correlation analysis between metabolite features and microbiota suggest that several bacterial taxa are associated with altered lipid and energy metabolism in PD.

## INTRODUCTION

Parkinson’ disease (PD) is the second most common neurodegenerative disorder and is associated with prominent gastrointestinal pathophysiological changes and symptoms^1^. Compositional alterations of gut microbiota in PD have been robustly demonstrated across multiple cohorts^2,3^. However, the functional implications of these changes regarding microbiota-host interactions and PD pathology and progression are poorly understood. Alterations of fecal and serum metabolites and inflammatory markers have been described in relation to gut microbiota^4–6^, but findings have been inconsistent except for reproducible findings of reduced fecal short-chain fatty acid levels in PD^7^.

The Helsinki cohort has so far been analyzed for microbiome correlations with clinical features^2,8^ and disease progression^9^. We have also studied the oral and nasal microbe communities^10^. Recently, the immune response and fecal SFCA levels were studied among the same individuals^6^.

In the current study we aimed to characterize links between untargeted serum metabolomics, microbiota composition, and clinical symptoms in PD as compared to healthy control subjects.

## RESULTS

### Metabolome analysis

Data-driven metabolomics profiling of serum samples was undertaken, identifying a total of 7585 metabolite features. Support Vector Machines (SVM) using RBF kernel showed 81% classification accuracy using GC-MS metabolic profiles, and 77% and 72% classification accuracy was achieved using LC-MS positive and negative mode ionisation data, respectively (Table 1).

**Table 1:**
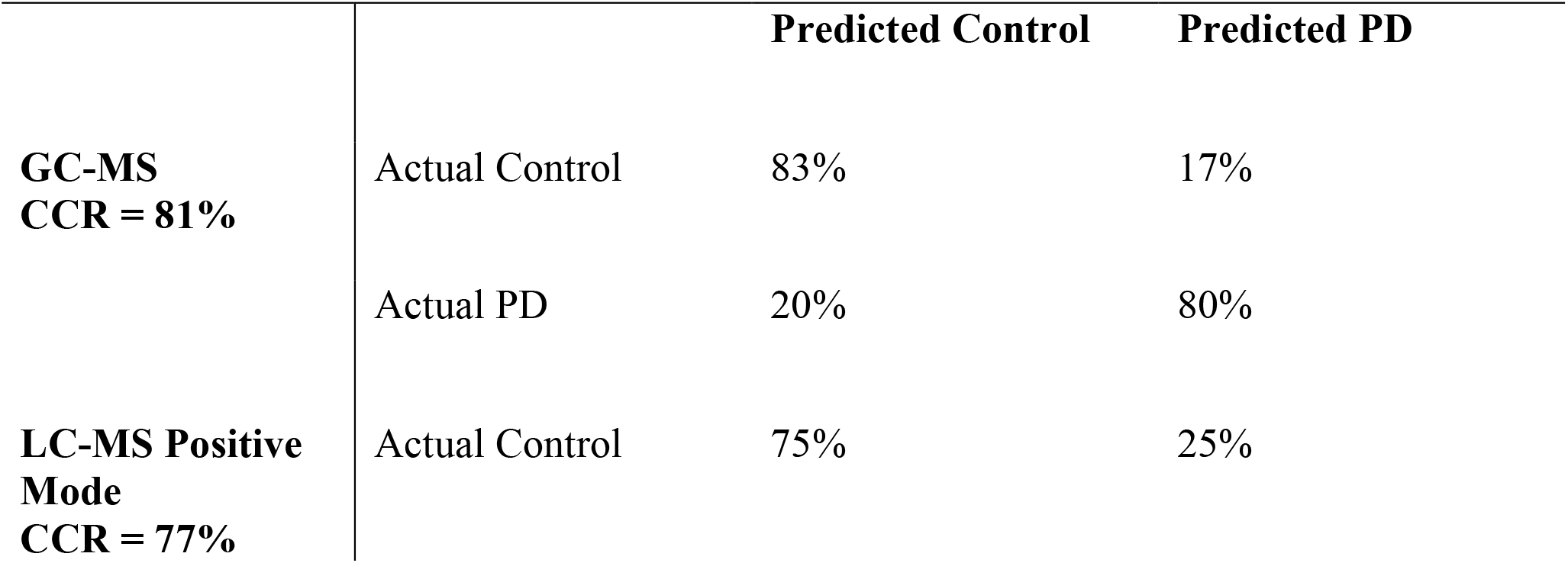
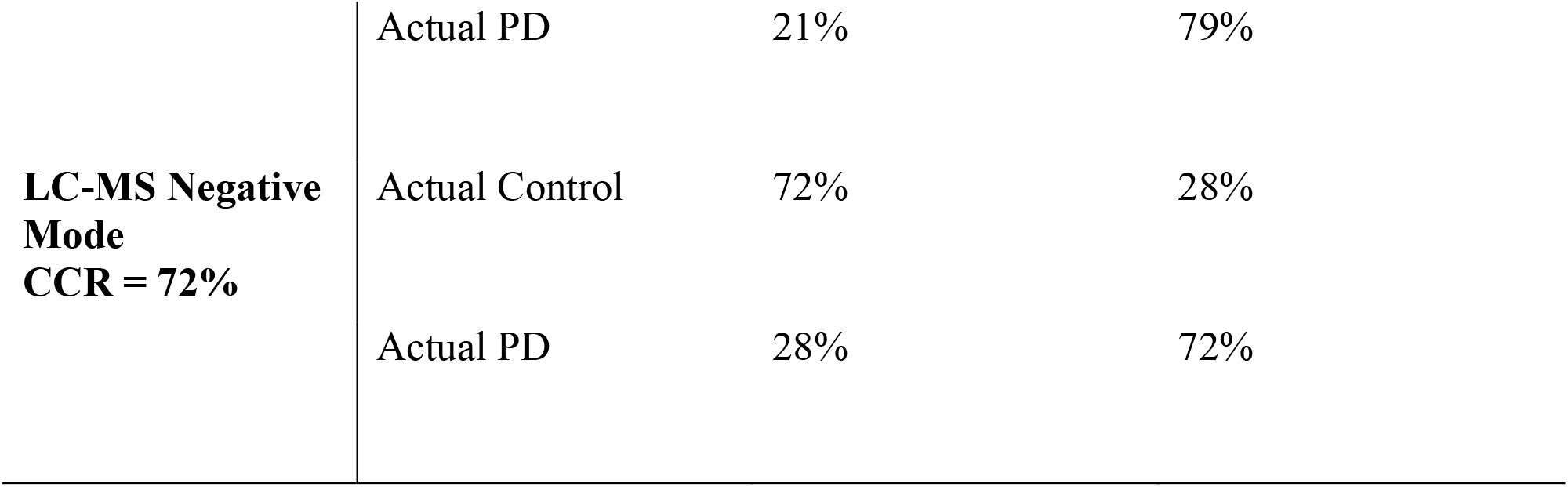
Confusion matrices for (a) LC-MS positive mode data, (b) LC-MS negative mode data, (c) GC-MS data. For each data, confusion matrix shows an average of 100 models tested by resampling. Each time 60% data were used as training set and 40% were used as test set. Average correct classification rate (CCR) is represented for each of the data. Upon permutation of class labels, LC-MS positive mode CCR dropped to 49%, LC-MS negative mode CCR dropped to 47% and GC-MS CCR dropped to 47%.

Variable selection of differentially abundant metabolite features between Controls and PD subjects was carried out using SVM recursive feature elimination to select the top 10% of variables from profiling experiments. A total of 139 features (i.e. metabolites) were selected: 101 features from LC-MS data and 38 features from GC-MS data (Table 2). Pathway enrichment analysis using all the 7585 metabolomics features revealed significant changes in carnitine shuttle, vitamin E metabolism, glycerophospholipids, sphingolipids, fatty acids and aminoacyl-tRNA biosynthesis amongst 20 perturbed pathways (Table 3). These features were also putatively annotated based on accurate mass match at 5ppm using human metabolome database (HMDB v.4)^11^ and LipidMaps^12^ following Metabolite Standards Initiative (MSI)^13^ at Level 3. Age, gender, BMI, dietary components, and other clinical variables did not show any evidence of confounding effects on the selected 139 metabolites. There was no effect of hypercholesterolaemia status on classification between PD and controls (Table 4). Within the PD group, no association was established between clinical variables and all 7585 metabolite features after adjusting for time since motor symptom onset, age at sampling, and other known covariates.

**Table 2:**
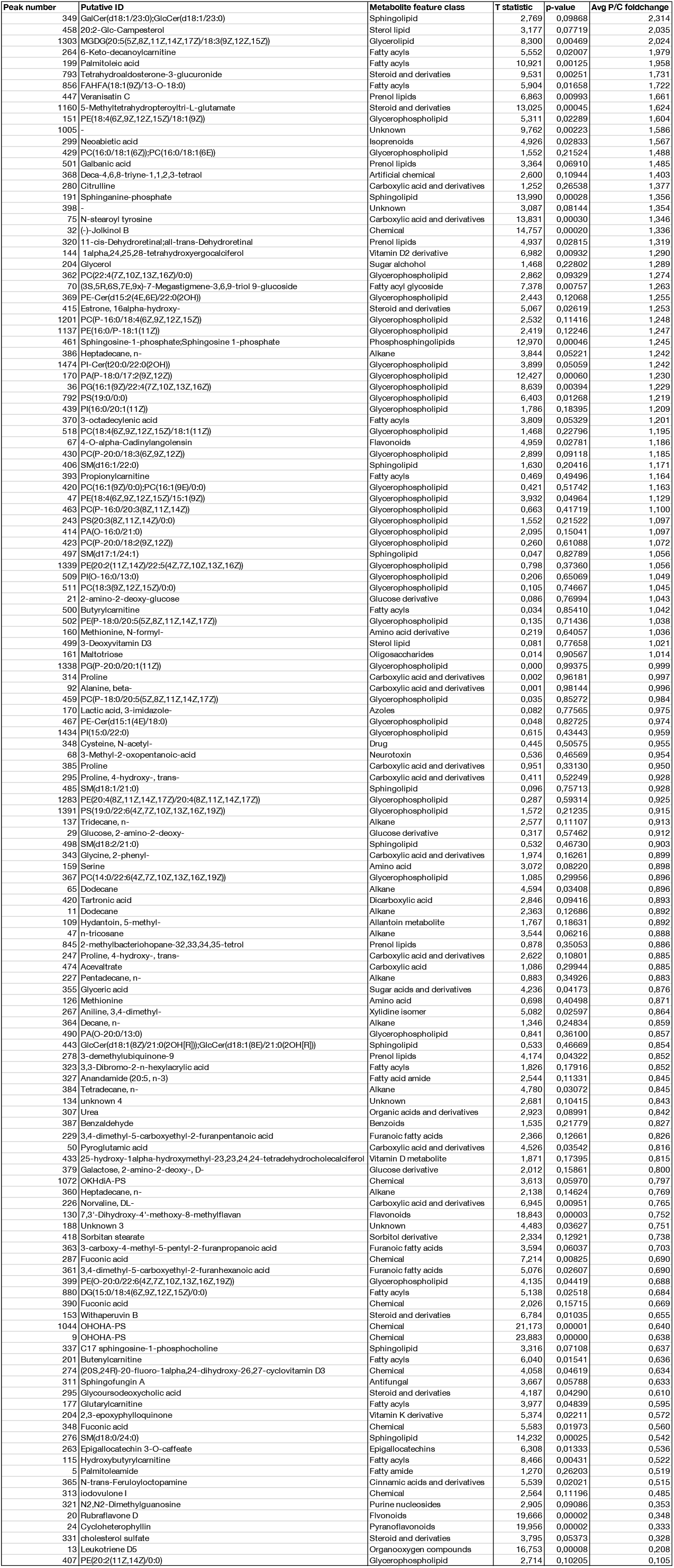
Differentially abundant metabolite features between PD and Controls, organized in descending order of effect size.

**Table 3:** Results from pathway analysis for LC-MS and GC-MS data. Metabolite overlap shows the number of metabolites that overlap on the total metabolites on pathway indicated by pathway size.

| Analytical Platform | Pathway name | Metabolite overlap | Pathway size | p-value |
| --- | --- | --- | --- | --- |
| LC-MS (pos mode) | Carnitine shuttle | 18 | 27 | 0.00934 |
|  | Vitamin E metabolism | 18 | 34 | 0.01417 |
|  | Glycosphingolipid metabolism | 15 | 28 | 0.01585 |
|  | N-Glycan Degradation | 5 | 6 | 0.01624 |
|  | Porphyrin metabolism | 13 | 25 | 0.0209 |
|  | Glycerophospholipid metabolism | 15 | 31 | 0.02649 |
|  | Saturated fatty acids beta-oxidation | 8 | 15 | 0.03312 |
|  | Linoleate metabolism | 9 | 18 | 0.03892 |
|  | Squalene and cholesterol biosynthesis | 17 | 39 | 0.04848 |
| LC-MS (neg mode) | De novo fatty acid biosynthesis | 13 | 18 | 0.00192 |
|  | Fatty acid activation | 12 | 17 | 0.00209 |
|  | Hexose phosphorylation | 6 | 7 | 0.00292 |
|  | Glycosphingolipid metabolism | 10 | 16 | 0.00384 |
|  | Caffeine metabolism | 6 | 10 | 0.01308 |
|  | Phosphatidylinositol phosphate metabolism | 4 | 6 | 0.02081 |
|  | Fructose and mannose metabolism | 4 | 6 | 0.02081 |
|  | Fatty Acid Metabolism | 4 | 6 | 0.02081 |
|  | Starch and Sucrose Metabolism | 3 | 4 | 0.03001 |
|  | Glycerophospholipid metabolism | 10 | 22 | 0.03784 |
| GC-MS | Aminoacyl-tRNA biosynthesis | 6 | 48 | 0.00011601 |
|  | Pantothenate and CoA biosynthesis | 3 | 19 | 0.0037957 |
|  | Valine, leucine and isoleucine biosynthesis | 2 | 8 | 0.007673 |
|  | Phenylalanine metabolism | 2 | 10 | 0.012069 |

**Table 4:** Confusion matrix generated from PLS-DA model using all the features from metabolomics data in a single model shows correct classification rate of 82%. The PLS-DA model of all metabolomics features adjusted for age, BMI, gender, dietary and clinical variables had correct classification accuracy of 84% (a gain of 2%) indicating the contribution of clinical variables does not significantly affect the classification for PD. Similarly, PLS-DA model generated for all metabolomics features, adjusting for hypercholesterolaemia does not improve much (0% gain) indicating the contribution of hypercholesterolaemia does not have an effect in classification for PD.

| Model |  | Predicted Control | Predicted PD |
| --- | --- | --- | --- |
| All metabolomics data, CCR = 82% | Actual Control | 79% | 21% |
|  | Actual PD | 15% | 85% |
| All metabolomics data adjusted for age, BMI, gender, dietary and clinical features, CCR = 84% | Actual Control | 82% | 18% |
|  | Actual PD | 14% | 86% |
| All metabolomics data adjusted for hypercholesterolaemia, CCR = 82% | Actual Control | 85% | 15% |
|  | Actual PD | 21% | 79% |

### Metabolome-microbiome correlation analysis

Correlation analysis between the selected 139 metabolite features and bacterial taxa at genus, family, and phylum levels was performed separately for the PD and Control groups to facilitate their contrast (Supplementary Tables 1-6). All results’ tables have been curated to obtain the best possible putative identification of the metabolite peak IDs at MSI level 3 identification level. Tables 2 and 5, as well as the two genus-level supplementary tables (Supplementary Tables 1 and 2) contain a metabolite “class” identification column (see Methods section for details). All supplementary tables contain all identified taxa/metabolite correlation pairs (using the selected 139 metabolite features and all bacterial abundance data) that show posterior mean correlation values at or above 0.3 and at a 95% “confidence level” (see Methods section for more details).

**Table 5:** Selected results based on taxa previously reported in the literature at genus, family, and phylum levels. Red numbers represent negative correlations, and Blue positive correlations. The last column presents a consensus on direction of effect based on previous reports: before the slash (/), we report the result obtained in Aho *et al*. (2019)^9^ using the same bacterial data as in the present study; after the slash, we report the consensus reported by Boertien *et al*. (2020)^3^ (see that study for details). NA means that no result for that taxon is available in that study^9^.

| Bacterial Genus: | Metabolite Peak ID | Metabolite MSI ID | Class | p2.5 | post. mean corr. | p97.5 | Taxon Diff Abund Direction Consensus<br>(Velma et al. 2019 / Jeffrey et al. 2020) |
| --- | --- | --- | --- | --- | --- | --- | --- |
| **Within-PD analysis:** |  |  |  |  |  |  |  |
| Prevotella | X7253 | Proline | Carboxylic acid and derivatives | -0.5146 | -0.3307 | -0.1186 | decreased in PD / same |
| Prevotella | X7287 | Tartronic acid | Dicarboxylic acid | -0.5025 | -0.3148 | -0.1066 | decreased in PD / same |
| Prevotella | X7206 | N-formyl-methionine | Carboxylic acid and derivatives | -0.4957 | -0.3063 | -0.0929 | decreased in PD / same |
| Roseburia | X499 | 3-Deoxyvitamin D3 | Sterol lipid | -0.5249 | -0.3348 | -0.1159 | decreased in PD / same |
| Roseburia | X423 | PC(P-20:0/18:2(9Z,12Z)) | Glycerophospholipid | 0.1034 | 0.3070 | 0.4970 | decreased in PD / same |
| Roseburia | X370 | 3-octadecylenic acid | Fatty acyls | 0.1231 | 0.3300 | 0.5208 | decreased in PD / same |
| Lactobacillus | X6092 | 3,4-dimethyl-5-carboxyethyl-2-furanpentanoic acid | Furanic fatty acids | -0.5270 | -0.3246 | -0.0991 | increased in PD / mostly increased in PD |
| Lactobacillus | X7038 | PE(20:2(11Z,14Z)/22:5(4Z,7Z,10Z,13Z,16Z)) | Glycerophospholipid | 0.0916 | 0.3260 | 0.5291 | increased in PD / mostly increased in PD |
| Akkermansia | X7038 | PE(20:2(11Z,14Z)/22:5(4Z,7Z,10Z,13Z,16Z)) | Glycerophospholipid | -0.5454 | -0.3572 | -0.1350 | NA / increased in PD |
| Akkermansia | X6564 | PS(19:0/0:0) | Glycerophospholipid | 0.1056 | 0.3305 | 0.5304 | NA / increased in PD |
| **Within-Controls analysis:** |  |  |  |  |  |  |  |
| Bifidobacterium | X7163 | 2-amino-2-deoxy-glucose | Hexoses | -0.5043 | -0.3109 | -0.0854 | increased in PD / mostly increased in PD |
| **Bacterial Family:** |  |  |  |  |  |  |  |
| **Within-PD analysis:** |  |  |  |  |  |  |  |
| Bifidobacteriaceae | X7145 | PI-Cer(t20:0/22:0(2OH)) | Glycerophospholipid | 0.0948 | 0.3069 | 0.4977 | increased in PD / mostly increased in PD |
| Pasteurellaceae | X6860 | PE(16:0/P-18:1(11Z)) | Glycerophospholipid | 0.1106 | 0.3521 | 0.5620 | decreased in PD / NA |
| Lactobacillaceae | X32 | (-)-Jolkinol B | Chemical | -0.5073 | -0.3027 | -0.0664 | increased in PD / mostly increased in PD |
| Lactobacillaceae | X7038 | PE(20:2(11Z,14Z)/22:5(4Z,7Z,10Z,13Z,16Z)) | Glycerophospholipid | 0.0823 | 0.3074 | 0.5096 | increased in PD / mostly increased in PD |
| Verrucomicrobiaceae | X7038 | PE(20:2(11Z,14Z)/22:5(4Z,7Z,10Z,13Z,16Z)) | Glycerophospholipid | -0.5430 | -0.3579 | -0.1496 | NA / increased in PD |
| Verrucomicrobiaceae | X6564 | PS(19:0/0:0) | Glycerophospholipid | 0.1288 | 0.3413 | 0.5297 | NA / increased in PD |
| Erysipelotrichaceae | X7253 | Proline | Carboxylic acid or derivative | 0.1616 | 0.3715 | 0.5648 | NA / mostly increased in PD |
| **Within-Controls analysis:** |  |  |  |  |  |  |  |
| Bifidobacteriaceae | X7163 | 2-amino-2-deoxy-glucose | Hexoses | -0.5127 | -0.3171 | -0.1049 | increased in PD / mostly increased in PD |
| Lactobacillaceae | X7183 | Beta-alanine | Carboxylic acid | -0.5079 | -0.3062 | -0.0822 | increased in PD / mostly increased in PD |
| Lactobacillaceae | X7081 | PS(19:0/22:6(4Z,7Z,10Z,13Z,16Z,19Z)) | Glycerophospholipid | 0.1223 | 0.3725 | 0.5795 | increased in PD / mostly increased in PD |
| Enterobacteriaceae | X485 | SM(d18:1/21:0) | Sphingolipid | 0.1140 | 0.3280 | 0.5281 | NA / mostly increased in PD |
| Enterobacteriaceae | X430 | PC(P-20:0/18:3(6Z,9Z,12Z)) | Glycerophospholipid | 0.1292 | 0.3299 | 0.5325 | NA / mostly increased in PD |
| **Bacterial Phylum:** |  |  |  |  |  |  |  |
| **Within-PD analysis:** |  |  |  |  |  |  |  |
| Verrucomicrobia | X7038 | PE(20:2(11Z,14Z)/22:5(4Z,7Z,10Z,13Z,16Z)) | Glycerophospholipid | -0.5306 | -0.3414 | -0.1231 | NA / increased in PD |
| Verrucomicrobia | X6564 | PS(19:0/0:0) | Glycerophospholipid | 0.1064 | 0.3345 | 0.5302 | NA / increased in PD |

The within-PD analysis at genus level identified a total of 176 correlation pairs, while within-Controls analysis produced 202 pairs (Supplementary Tables 1 and 2, respectively). As can be seen, there is some overlap in the taxa and metabolites represented in the two groups, but overall there are substantial differences in the bacterial taxon-metabolite pair associations. To aid in the identification of possible links between metabolite classes and bacteria, we produced network figures for both within-PD and within-Controls results at genus level, using metabolite classes. (Figures 1 and 2). At family level, the within-PD analysis identified a total of 67 correlation pairs, while the within-Controls analysis identified a total of 85 pairs (Supplementary Tables 3 and 4, respectively). Finally, the within-PD analysis at phylum level identified a total of 17 correlation pairs and the within-Controls analysis identified a total of 6 pairs (Supplementary Tables 5 and 6, respectively).

**Figure 1:**
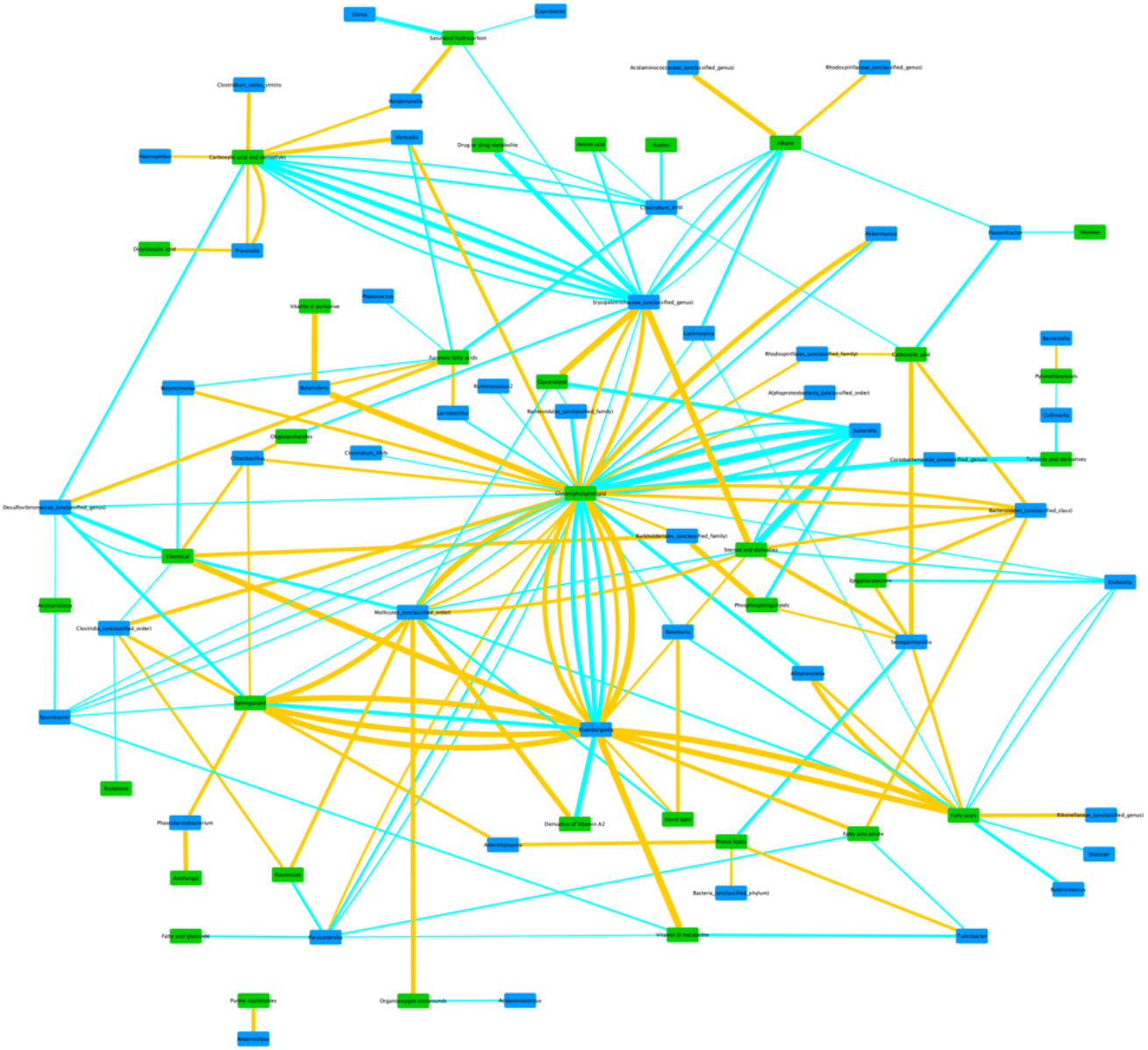
Network of within-PD correlations between bacterial genera and metabolite classes. Taxon groups unclassified at genus level may contain more than one genus (or higher taxon) in the same node, but were kept in the network for visualization purposes and to match the raw results from Supplementary table 1. Edge thickness represents the strength of the correlation. Blue edges represent positive correlations, and Orange edges represent negative correlations. Green nodes represent metabolite classes, and Blue nodes represent bacterial taxa.

**Figure 2:**
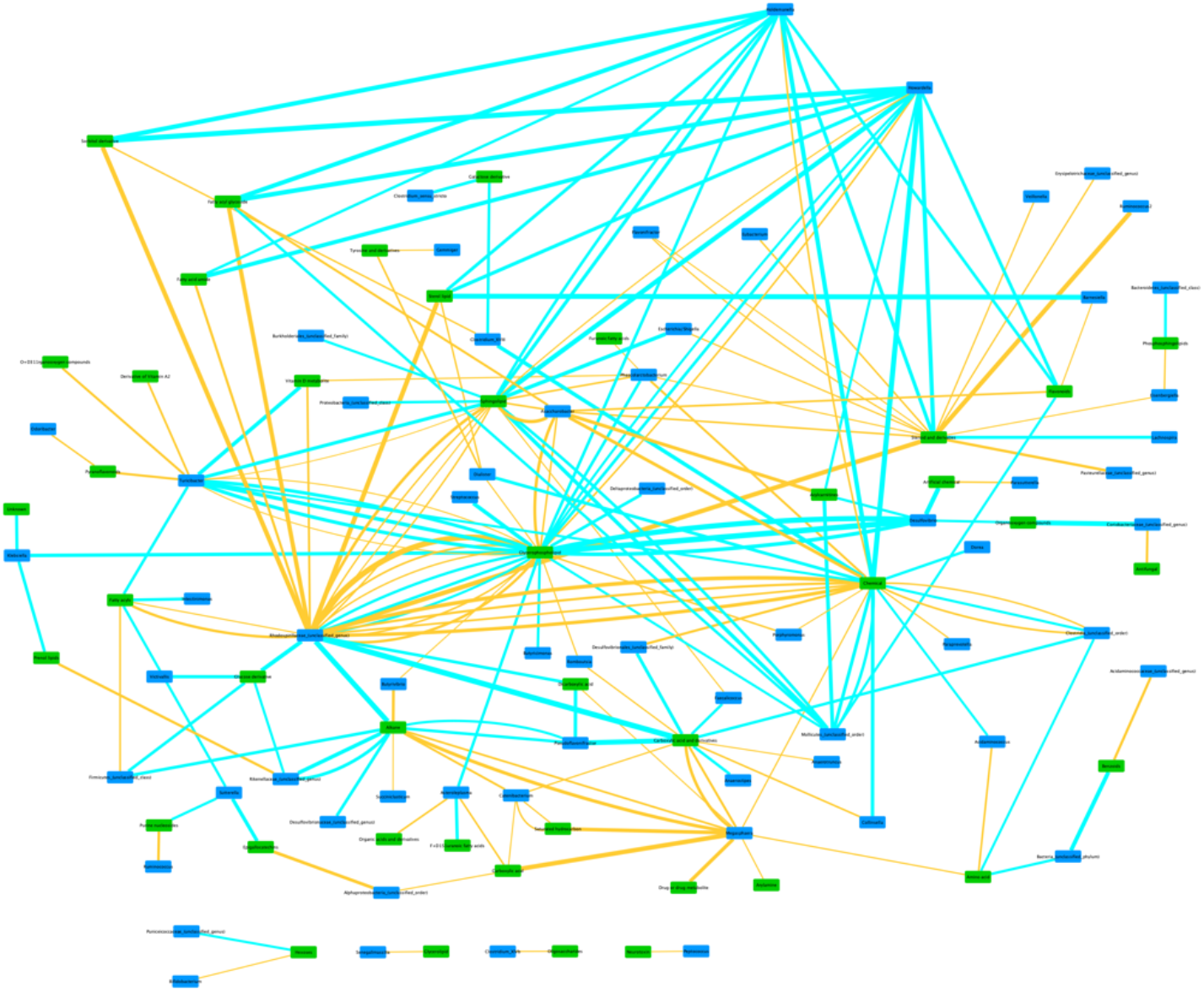
Network of within-Controls’ correlations between bacterial genera and metabolite classes. Taxon groups unclassified at genus level may contain more than one genus (or higher taxon) in the same node, but were kept in the network for visualization purposes and to match the raw results from Supplementary table 2. Edge thickness represents the strength of the correlation. Blue edges represent positive correlations, and Orange edges represent negative correlations. Green nodes represent metabolite classes, and Blue nodes represent bacterial taxa.

To further focus the study, we trimmed the correlation pairs down to only those containing bacterial taxa that were i) not unclassified at the target taxonomic level, ii) differentially abundant between PD and Control groups at one or both of the two time points of sample collection in a previous study using the same subject data^9^, and iii) taxa that were systematically reported previously in the PD microbiome literature as being differentially abundant between PD and Control groups^3,9^ (Table 5). The bacterial abundance data used in the present article corresponds to the second time point of sample collection in Aho *et al*. (2019)^9^. To aid in visualizing the relationships, a third within-PD network figure was produced, using genus-level data and metabolite classes as before, but limited to the trimmed correlation pairs (Figure 3).

**Figure 3:**
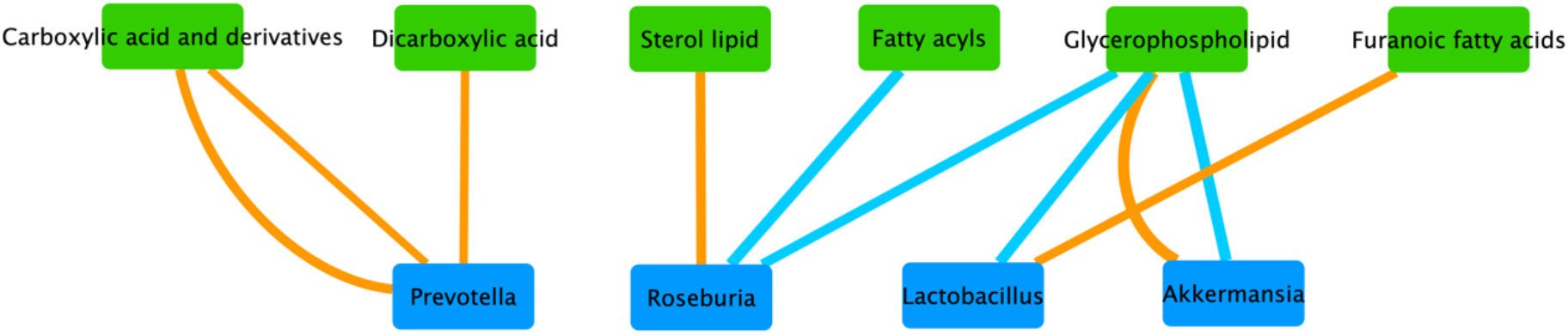
Network of within-PD correlations between bacterial genera and metabolite classes using selected results (Table 5). Edge thickness represents the strength of the correlation. Blue edges represent positive correlations, and Orange edges represent negative correlations. Green nodes represent metabolite classes, and Blue nodes represent bacterial taxa.

In the Helsinki cohort, six bacterial genera were previously reported as being differentially abundant (selected for the present article at an alpha threshold of statistical significance of 0.05 from the original 0.1), using one or more statistical methods^9^, between Control and PD groups at one or both time points, namely: *Bifidobacterium, Roseburia, Prevotella, Blautia, Lactobacillus*, and *Clostridium XIVa*. All these genera, except for *Clostridium XIVa*, have also been reported as being differentially abundant in previous publications contrasting a control group to PD patients^9^. Of these, *Prevotella, Bifidobacterium, Roseburia*, and *Lactobacillus* are also found to be correlated with one or more metabolite features in our genus-level analysis (Table 5).

Other differentially abundant bacterial genera reported previously in the PD microbiome literature^3^ besides those referred to in Aho *et al*. (2019)^9^ have also been found covarying with metabolite features in our dataset, and we also used that information for the purpose of focusing our study’s results. At genus level, *Akkermansia, Bifidobacterium, Faecalibacterium, Prevotella, Lactobacillus*, and *Roseburia* were reported multiple times in the literature, with only *Akkermansia* and *Faecalibacterium* not being reported in the Aho *et al*. (2019)^9^ study as being differentially abundant (Table 5).

In the Aho *et al*. (2019)^9^ study, seven bacterial families were reported as being differentially abundant between PD and Control groups at one or both time points, namely: *Bifidobacteriaceae, Prevotellaceae, Rikenellaceae, Lachnospiraceae, Pasteurellaceae, Lactobacillaceae*, and *Puniceicoccaceae*. All these families, except for *Puniceicoccaceae*, have also been reported as being differentially abundant in previous publications contrasting a control group to PD patients^9^. Three of them showed correlations with one or more metabolite features in our family-level analysis (Table 5). Boertien *et al*. (2019)^3^ also reported on the bacterial families most commonly found to be differentially abundant, namely *Bifidobacteriaceae, Prevotellaceae, Lachnospiraceae, Lactobacillaceae, Verrucomicrobiaceae, Enterobacteriaceae, Erysipelotrichaceae*, and *Ruminococcaceae*, with the last four families not being found to be differentially abundant at any time point in our cohort^9^ (Table 5).

For this study, we have also analysed our data at phylum level, unlike in Aho *et al*. (2019)^9^, and found correlations between various metabolites and the phyla *Lentisphaerae, Verrucomicrobia, Synergistetes*, and *Tenericutes* (Supplementary Tables 5 and 6). Some phyla recurrently found to be differentially abundant between PD and Control groups are *Verrucomicrobia, Firmicutes*, and *Bacteroidetes*^3^, although of those reported only *Verrucomicrobia* yielded correlations with metabolite features in our analysis (Table 5).

## DISCUSSION

### Serum Metabolome

Using mummichog approach, we have shown in this study that the identified serum metabolome differences in PD have functional significance on over 20 pathways, including carnitine shuttle, vitamin E metabolism, glycerophospholipids, sphingolipids, fatty acids, and aminoacyl-tRNA biosynthesis. In a separate study, we have demonstrated that carnitine shuttle, sphingolipids, and fatty acids pathways change in Parkinson’s sebum - these are within the 20 pathways enlisted above detected in serum in this study^14^. Energy metabolism is highly regulated by facilitation of long chain fatty acid β-oxidation. Also in frail^15^ elderly participants without Parkinson’s, dysregulation of carnitine shuttle and vitamin E metabolism was observed when compared to similarly aged resilient individuals^16^. Thus, perturbation of carnitine shuttle and vitamin E, along with fatty acids in serum metabolome may indicate a significant change in energy metabolism during PD. This can further be supported by changes observed in sphingolipid metabolism, a key pathway in cell signalling and regulation.

Dysregulation of sphingolipids is known to be associated with α-synucleinopathy^17,18^, changes in lysosomal metabolism, and in mitochondrial metabolism observed in PD^19,20^. Decreased long-chain acylcarnitines due to insufficient β-oxidation has been shown to carry potential for early diagnosis of Parkinson’s^21^, especially 12-14 long chain acylcarnitines. In recent work studying the gut microbiome, Rosario *et al*. (2021)^22^ have shown the role of bacterial folate and homocysteine metabolism in PD. Higher numbers of bacterial mucin and host degradation enzymes were linked to the manifestation of PD. The contribution of bacterial folate metabolism to human metabolic regulation is not entirely clear. Folate is an essential vitamin B, that maintains methylation reactions. The liver, via many methylation reactions in post-translational modifications, regulates the synthesis of hormones, creatine, carnitine, and phosphatidylcholine^23^. If methylation capacity is compromised due to an alteration in folate metabolism, there may be impaired phosphatidylcholine synthesis along with shunted or disrupted carnitine shuttle observed in our results. Altered carnitine metabolism, fatty acids, and steroid metabolism was also observed in a metabolomics profiling study recently reported^24^. Several studies have reported decreased levels of carnitine and acylcarnitines in plasma from PD patients^21,25,26^; however, according to Jiménez-Jiménez *et al*. (1997)^27^ no changes were observed in acylcarnitine levels in plasma or cerebrospinal fluids of PD participants. Thus, there is no clear evidence of direction in which carnitines are expressed but there is much research evidence that suggests a link between perturbations in carnitine shuttle owing to protective mechanism of acylcarnitines leading to changes in other fatty acids and eventually the lipid make-up in PD.

### Correlations between bacterial taxa and metabolites features

Regarding the correlations between bacterial taxa and metabolite features, and given that metabolite MSI 3 ID is a putative identification, we will mostly focus the following discussion at the level of metabolite classes. When mentioning specific bacterial taxa in terms of correlation results, we will report between brackets if the taxon is always (or usually) reported in the literature as being over- or underrepresented in PD (see the last column in the Table 5 for details), as well as the signal of the correlation detected in our study.

At all bacterial taxonomic levels investigated in this study, the most commonly detected correlations were found with putative metabolites in the glycerophospholipid class and with lipids in general. This is the case both in our non-trimmed results (Supplementary Tables 1-6) and in the trimmed results focusing only on bacterial taxa found in the previous literature as being differentially abundant between Controls and PD cases (Table 5). Specifically, in the within-PD analysis and focusing the discussion on the trimmed results, we find correlations between various glycerophospholipids and *Roseburia* (decreased in PD; positive correlation), *Lactobacillus* (increased in PD; positive correlation), *Akkermansia* (increased in PD; one positive and one negative correlation), *Bifidobacteriaceae* (increased in PD; positive correlation), *Pasteurellaceae* (decreased in PD; positive correlation), *Lactobacillaceae* (increased in PD; positive correlation), *Verrucomicrobiaceae* (increased in PD; one positive and one negative correlation), and *Verrucomicrobia* (increased in PD; one positive and one negative correlation) (Table 5). *Akkermansia, Verrucomicrobiaceae*, and *Verrucomicrobia* share the same positive and negative correlations with two metabolite features. *Lactobacillus* and *Lactobacillaceae* correlate positively with the same metabolite feature.

With the exception of *Roseburia* and *Pasteurellaceae*, all these taxa are usually found to be overrepresented in PD, and are mostly positively correlated with various glycerophospholipids, which are also found in our analysis to be mostly overrepresented in PD (Table 2). This is not the case in the within-controls analysis, in which the only detected correlations with glycerophospholipids are with *Lactobacillaceae* (positive correlation with a different metabolite) and *Enterobacteriaceae* (also a positive correlation with a different metabolite) (Table 5). The genus-level network figures for PD and Controls (Figures 1 and 2) also indicate the existence of possible alterations in bacterial metabolic dynamics in PD. Overall, these results suggest that these bacterial taxa, which have been found to be overabundant in PD in several studies, may be associated for the most part with an increase in glycerophospholipid abundance in PD.

All of these glycerophospholipids have an endogenous (human host) origin and are linked to cell signalling, lipid peroxidation, and lipid metabolism. These phospholipids are the main component of cell membranes in all known living systems, and play roles in various biological processes, including signal induction and acting as transporters. Interestingly, there are several genetic factors directly or indirectly related to glycerophospholipid metabolism, such as *PLA2G6*^17^, that are associated with PD risk (*PLA2G6* is the cause of early-onset PARK14-linked dystonia-parkinsonism^28,29^). Alpha-synuclein, characteristically found in aggregates within Lewy bodies in the brains of PD patients, directly binds to negatively charged phospholipids in the cells’ lipid membranes, and exhibits preferential binding to small lipid vesicles^30^. The binding of alpha-synuclein to lipid membranes can also lead to alterations in their bilayer structure that can induce the formation of those lipid vesicles^31^. Very importantly in the PD context, these interactions between lipid membranes and alpha-synuclein affect its rate of aggregation, and can lead to disruption of membrane integrity both *in vitro* and *in vivo*^32^. It has also been shown that the association of soluble alpha-synuclein with planar lipid bilayers results in the formation of aggregates and small fibrils^33^. Exposure to docosahexaenoic acid (DHA), which accounts for 60% of glycerophospholipid esterified fatty acids in the plasma membrane, gradually assembles alpha-synuclein into amyloid-like fibrils, with the notable feature that DHA itself becomes part of the aggregate^34^. Notably, alpha-synuclein gene expression is increased with elevated DHA intake, and the resulting oligomers are toxic to cells^35,36^. Alpha-synuclein also binds with specific phospholipids in mitochondrial membranes, modulating the efficiency of mitochondrial energy production^37^, with various mitochondrial phospholipids appearing to have an effect on alpha-synuclein toxicity^17^. Thus, the interaction between alpha-synuclein and various phospholipids and their metabolism may play an important role in PD pathogenesis, and gut microbiota may be implicated in these interactions.

We also detected correlations with other lipids in the within-PD analysis. *Roseburia* (decreased in PD) negatively correlates with a sterol lipid, probably of dietary origin. *Roseburia* is also positively correlated with a metabolite in the fatty acyl class, possibly also associated with diet. *Lactobacillus* (increased in PD) is negatively correlated with a furanoic fatty acid, which is associated with cell signalling, lipid peroxidation, lipid metabolism, and lipid transport metabolism, with dietary, human, and/or bacterial origin. Half of the metabolites from the fatty acyl class were found to be overrepresented in PD in the selected 139 metabolite features in our blood serum data (Table 2), but overall underrepresented in the PD sebum data from Sinclair *et al*. (2021)^14^.

In the within-controls analysis we detect a positive correlation between *Enterobacteriaceae* (increased in PD) and a sphingolipid, which is associated with membrane stabilization, lipid peroxidation, and lipid metabolism, of endogenous origin. In our study, this metabolite feature is slightly decreased in PD (Table 2) and no correlation is found between it and PD-linked bacterial taxa in the within-PD analysis.

Although the interpretation of these results regarding lipids in general is not suggestive of a particular pattern as in the case of glycerophospholipids, it is nevertheless interesting that virtually all correlations, positive or negative, with lipids are detected in the PD group, with most lipids detected in our study being overrepresented in PD relative to the control group (Table 2). Also interesting is that the majority of the identified correlations between metabolite features and the trimmed bacterial taxa list are with lipids, with relatively few other metabolite groups represented in the results (Table 5). The importance of this link between lipid metabolism and PD can’t be overstated: as mentioned earlier in the context of phospholipids, recent research shows that alpha-synuclein binds preferentially to specific lipid families and molecules, and that the latter promote alpha-synuclein interaction with synaptic membranes and affect alpha-synuclein oligomerization and aggregation. These same lipid-protein complexes also affect lipid metabolism by interfering with the catalytic activity of lipid enzymes in the cytoplasm and lipases in lysosomes. Lipid compositional alterations in PD have also been reported in brain and plasma, as well as linked to oxidative stress, inflammation, and progressive neurodegeneration through pro-inflammatory lipid mediators (see Alecu *et al*. (2019)^17^ for a full review on the role of lipids in PD). The link between lipids and bacteria in PD, if any, would probably consist of the bacterial modulation of lipid intake through diet and its differential effect on the bioavailability of those lipids in the host. The results of our study, by establishing associations between bacterial taxa found to be differentially abundant between Controls and the PD group and lipid metabolites present in serum that are themselves differentially abundant between both groups, suggests that such a scenario could have a role in PD pathology and development.

Further correlations with putative metabolites in other classes have also been detected in our study, in particular in the hexoses class and carboxylic acid or derivatives class. In the hexoses case, two negative correlations for the same metabolite feature were found for *Bifidobacterium* and *Bifidobacteriaceae* (both taxon levels increased in PD; Table 5). These two correlations are only found in the within-controls analysis. The metabolite is probably endogenous in origin and is involved in sugar metabolism shunts, diverting a proportion of glucose from the main glycolytic path and returning metabolites at the level of triose phosphate and fructose 6-phosphate.

Finally, a positive correlation in the within-PD analysis is found between a metabolite feature in the carboxylic acid or derivatives class and the bacterial family *Erysipelotrichaceae*, which is mostly found in previous studies to be overrepresented in PD. This metabolite feature is tentatively identified as proline and may have a microbiome or endogenous source. Four different metabolite features found to be differentially abundant in our data are tentatively identified as proline (Table 2), and in all cases show a slight decrease in mean abundance in PD. Interestingly, L-proline can act as a weak agonist for glycine and glutamate receptors^38^, like NMDA, AMPA, and kainite. Both glutamate and glycine are neurotransmitters. Although that is not the case in our data, proline has been reported previously as being overrepresented in PD^24^ and is known to be linked to protein metabolism and structure, cell differentiation, conceptus growth and development^39^, and gut microbiota community re-equilibration in cases of dysbiosis, with L-proline dietary supplementation being known to affect gut microbial composition and gut concentrations of several bacterial metabolites^40^. One of the detected correlations with *Prevotella* also involves (putative) proline. *Prevotella* and *Prevotellaceae*, when detected in PD studies, are usually found decreased in PD. In this study it correlates negatively with proline, which is found to be slightly underrepresented in the PD group.

## METHODS

### Study subjects, clinical data, and sampling

The present study uses bacterial abundance data that was used in a previously published study by Aho *et al*. (2019)^9^. The study subjects and associated clinical data in the present study is similar to the data referred to previously in that study as “follow-up” timepoint, with minor changes specific to the present study: of the original 128 subjects, 61 control subjects and 63 PD patients were used in the present study, i.e. 3 control subjects and 1 PD subject less than in the original cohort (C75, C82, C123, and P119). This difference in sample numbers was due to insufficient metabolite data available to perform the study.

For DNA sequencing, the stool samples were collected and stored as described earlier^9^. For serum samples, blood was drawn at the study visit and after processing immediately transferred to −20°C and subsequently to −80°C. Samples were shipped overnight on dry ice from Helsinki to Manchester for analysis.

### Sample preparation and metabolomics methods

#### Metabolomics sample preparation

Untargeted metabolite profiling was performed on serum samples that were collected from participants stored at −80 °C prior to analysis. Complementary coverage of metabolites was obtained using Ultra-High Performance Liquid Chromatography Mass Spectrometry (UHPLC-MS) and gas chromatography mass spectrometry (GC-MS). The procedures were adapted from the Dunn^41^ and Begley^42^ protocols as summarized here:

Metabolites were extracted from the serum samples by individually adding 400 µL of cold methanol to 200 µL of serum. This was followed vortexing and centrifugation (17,500 × *g*) to yield a metabolite rich supernatant that was split into two aliquots and lyophilised for 12 h.

Resultant metabolite pellet was stored at −80 °C until analysis. A pooled QC standard was also generated by combining 20 µL aliquots of each sample into a pooled vial with subsequent 200 µL aliquots from the pool, being extracted identical to each sample.

#### LC-MS method parameters

Processed metabolite pellets were defrosted at 4 °C and subsequently reconstituted in 100 µL of 95:5 H2O:MeOH (*v/v)*. UHPLC-MS analysis was performed using an Accela UHPLC with cooled auto sampler system coupled to an electrospray LTQ-Orbitrap XL hybrid mass spectrometer (ThermoFisher, Bremen, Germany). Analysis was carried out in positive and negative ESI modes while samples were completely randomised to negate for any bias. The mobile phases and gradient elution profile were as tabulated below:

| Time (min) | Mobile Phase A (95:5 H<sub>2</sub>O:MeOH with 0.1% Formic acid) composition | Mobile Phase B (95:5 MeOH:H<sub>2</sub>O with 0.1% Formic acid) composition |
| --- | --- | --- |
| 0 | 100 | 0 |
| 1 | 95 | 5 |
| 12 | 5 | 95 |
| 20 | 5 | 95 |
| 22 | 95 | 5 |
| 25 | 95 | 5 |

From each sample vial, 10 µL of the extract was injected onto a Hypersil GOLD UHPLC C18 column (length 100 mm, diameter 2.1 mm, particle size 1.9 µm, Thermo-Fisher Ltd. Hemel Hempsted, UK) held at a constant temperature of 50 °C with a solvent flow rate of 400 µL min^−1^.

Prior to analysis, LTQ-Orbitrap XL was calibrated according to manufacturer’s instructions using caffeine (20 µg mL^−1^), the tetrapeptide MRFA (1 µg ml^−1^) and Ultramark 1621 (0.001%) in an aqueous solution of acetonitrile (50%), methanol (25%) and acetic acid (1%). The data acquisition was performed in centroid mode with 30K mass resolution and scan rate of 400ms per scan. The masses were measured between 100-1200 *m/z* range with source gases set at sheath gas = 40 arb units, aux gas = 0 arb units, sweep gas = 5 arb units. The ESI source voltage was set to 3.5 V, and capillary ion transfer tube temperature set at 275 °C.

#### LC-MS data processing

Xcaliber software (v.3.0; Thermo-Fisher Ltd. Hemel Hempsted, U.K.) was used as the operating software for the Thermo LTQ-Orbitrap XL mass spectrometer. Data processing was initiated by the conversion of the standard UHPLC .*raw* files into the .*mzML* format using Proteowizard^43^. Subsequently, peak picking was carried out in RStudio^44^ using the XCMS^45^ package for data deconvolution (http://masspec.scripps.edu/xcms/xcms.php). The output data was a matrix of mass spectral features with accurate *m/z* and retention time pairs. Any missing values after deconvolution were replaced using k-nearest neighbours algorithm. Peaks that with relative standard deviation of more than 20% within pooled QCs were removed. The remaining data was normalised with total ion count to account for injection to injection signal variations, log10 transformed, and pareto scaled prior to statistical analysis.

#### GC-MS method parameters

Analysis of serum samples was also carried out on a Agilent 7250 GC-Time-of-Flight mass spectrometer coupled to a Gerstel-MPS autosampler. Two step derivatization of metabolite pellets thawed at 4 °C was carried out as described in the Begley protocol^42^. The source temperature was set to 230°C and quad temperature was at 150°C. The total run time was 25 minutes for 10 µL sample injected each time. The sample was injected in split mode with 20:1 split ratio and split flow of 20 mL per minute. Agilent CP8944 VF-5ms column was used for separation (30m x 250µm x 0.25µm). With a 5 minute solvent delay at the start of run, gradient elution method was used to elute and separate analytes from serum. The oven temperature was ramped from 70°C to 300°C with an increase of 14°C per minute. At 300°C the temperature was held for 4 minutes before dropping back to starting conditions.

#### GC-MS data processing

The raw data files were in Agilent. *D* format that were converted to .*mzML* format using Proteowizard^43^. Peak picking was carried out in RStudio^44^ using an in-house script for the eRah^46^ package for GC-MS peak picking and deconvolution. The peaks were annotated using eRah’s MassBank library. Any missing values after deconvolution were replaced using k-nearest neighbours algorithm. Peaks that with relative standard deviation of more than 20% within pooled QCs were removed. The remaining data was normalised with total ion count to account for injection to injection signal variations, log10 transformed, and pareto scaled prior to statistical analysis.

All metabolites successfully annotated within both the LC-MS and GC-MS analysis were assessed and scored at *MSI level 3 putative identification* according to rules set out by the Chemical Analysis Working Group of the Metabolite Standards Initiative^13^.

### Sample preparation and DNA sequencing

The stool sample preparation and subsequent sequencing of the bacterial 16S rRNA genes has been described earlier^9^. All details, as well as the results of the full analysis of the 16S rRNA gene amplicon sequence data can be obtained from that publication.

### Bioinformatics and Statistical Data Analysis

#### Statistical analysis of metabolomics data

In this untargeted profiling study, we performed

##### (i) SVM models

Using Python *via* Orange user interface, SVM models were generated for this analysis. The data were pre-treated as described earlier. The data were then split into train (60% data) and test (40% data), and resampling was repeated 100 times. The SVM model was generated with linear-kernel, cost (C) was set to 1.5, and regression loss epsilon was set at 0.10.

##### (ii) feature selection

To select metabolite features (i.e. features differentially abundant between the Control and PD groups) contributing to the SVM models, the mSVM-RFE algorithm was used^47^. The iterative algorithm worked backwards from an initial set of features consisting of all the variables (i.e. metabolite features) in the dataset. At each iterative round, firstly a simple linear SVM was fitted, then features were ranked based on their weights in the SVM solution and lastly, the algorithm eliminated the feature with the lowest weight. In order to stabilize these feature ranking, at each iteration cross validation resampling was used. By using k-fold cross validation (k=10) multiple SVM-RFE iterations were carried out. From the resultant ranked feature list, the top 10% of the features were selected for further interpretation as they contributed the most towards SVM models.

##### (iii) variable selection

In order to investigate the effect of 84 clinical and technical variables, these were used along with the metabolomics abundance data to create PLS-DA models. The effect of additional metadata variables was marginal, indicating no significant contributions by these variables in classification or prediction of phenotypes (Table 4). Correct classification rate as an average of 250 bootstraps was 84% compared to 82% obtained using only metabolomics data.

##### (iv) within-PD metabolite-clinical features associations

All metabolite features were regressed against clinical features of Parkinson’s *viz*. GDS-15^48^ (depression), MMSE^49^ (cognition), NMSS^50^ (non-motor symptoms), RBDSQ^51^ (REM-sleep behavior disorder), Rome III constipation score^52^, Rome III IBS status, Wexner score^53^ (constipation), SCS-PD^54^ (drooling), SDQ^55^ (dysphagia), UPDRS II-III, Hoeh & Yahr scale, and progression category from Aho *et al*. (2019)^9^, while adjusting for confounders. All RBDSQ and Progression categories were adjusted for age at sampling and time since motor onset. SCS-PD, SDQ, UPDRS-II, UPDRS-III and Hoehn & Yahr were also adjusted for LED. UPDRS-III was additionally adjusted for beta blockers. Wexner score and Rome III IBS status and Rome III constipation scores were also adjusted for anticholinergic medication, constipation medication, opioids, and tricyclic medications, as well as dietary fiber intake. SCS-PD was additionally adjusted for anticholinergic and tricyclics. GDS-15 was additionally adjusted for SSRI medications. MMSE was additionally adjusted for anticholinergic and tricyclic medications.

The features that had continuous scaled values were subjected to linear regression with elastic net penalty (50:50, L1:L2) and partial least squares regression with 20 PCs, for 1000 iterations. The features that had categorical values were subjected to support vector machines with RBF kernels (cost was set to 1.5 and regression loss epsilon was set at 0.10), random forest (10 trees), and logistic regression with ridge penalization with cost value of 1.

##### (v) pathway analysis

In this data driven approach, we have interrogated data generated from an untargeted profiling study. It is often impractical to identify each peak in a metabolomics profiling study as it could contain upwards to 5000 features in a single sample. To identify them accurately, the only option is to purchase commercial standards and perform MS/MS analysis in both samples and standards and then match fragmentation spectra. This could be relevant when performing a targeted analysis with a defined set of metabolites. Computationally predicted *m/z* based identification alone is not adequate for pathway analysis due to multiple metabolite matches to single *m/z*. Thus, we have employed mummichog analysis that does not depend on identification of metabolites and then mapping on pathways. Instead mummichog leverages the collective power in the organisation of metabolic networks. If a list of *m/z* values truly reflect a biological activity, the true metabolites that are represented by these *m/z* values should show enrichment on a local structure of a metabolic network. If the measured *m/z* matches to a falsely represented metabolite, the distribution will be observed randomly. The overall significance of mapping and pathway enrichment is estimated by ranking the p-values from the real data among the p-values from permutation data to adjust for type I error, along with penalisation. Thus, a robust functional metabolic network gives us insight into our data more than identifying a handful of features.

Mummichog^56^ (v.1.0.9) pathway analysis was used to predict network activity from pre-processed UHPLC-MS metabolomics data. The full metabolite data set consisting of 5897 and 1258 features from LC-MS positive and negative mode, respectively, was used as an input. Pathway enrichment analysis was performed on annotated 429 GC-MS features using MetaboAnalystR^57^.

#### Data pre-processing for 16S rRNA gene sequence data

The bioinformatics pipeline for the 16S rRNA gene amplicon sequence data used in the present study was the same as described in Aho *et al*. (2019)^9^ given that the same data was used for the present study minus the above mentioned 4 samples that were excluded from the present analysis during quality control of the metabolomics data. Please refer to the above study for details.

#### Metabolome-microbiome correlation analysis

Statistical data analysis specific to the 16S rRNA gene sequence data was performed and reported in full earlier^9^. Putative confounders (technical or otherwise) identified in that analysis were taken into account in the correlation modelling performed in the present paper (see below for details).

For correlation analysis between metabolites and bacterial taxa at genus, family, and phylum levels, we used the *fido*^58^ package (v.0.1.13; the package was formerly known as *stray*) for the R Statistical Programming Software^59^ (v.3.6.0). *fido* provides a framework for inferring multinomial logistic-normal models which can account for zeros and compositional constraints, as well as sampling and technical variation present in sequence count data^58^. For the present study we used the function *orthus* from *fido* which enables joint modelling of multivariate count data (e.g. 16S rRNA gene amplicon sequence data) and multivariate Gaussian data (e.g. metabolomic data on the log-scale).

For samples *j*∊{1, …,*N*} we denote by *Y*_*j*_ the observed *D*_1_-dimensional vector of sequence counts, *Z*_*j*_ the standardized (i.e. Z-transformed) and log_10_-transformed *P*-dimensional vector of observed metabolite abundances, and *X*_*j*_ a *Q*-dimensional vector of covariates. Using this notation, the *orthus* likelihood model is given by

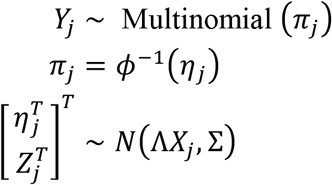

with priors Λ ~ *N*(Θ, Σ, Γ) and Σ ~ *Inverse Wishart* (Ξ, *v*) and with *ϕ*^−1^ denoting the inverse additive log-ratio (ALR) transform with respect to the *D*-th taxa^60^. Of note, the ultimate inference is invariant to the chosen ALR transform. This represents a joint linear model over the latent relative abundances of microbial taxa and metabolite abundances. For computational scalability this model was inferred using the multinomial-Dirichlet Bootstrap approximation to the marginal posterior density *p*(π | *Y*) that is available in *fido*. The multinomial-Dirichlet bootstrap approximates the true marginal posterior density using the posterior of a Bayesian multinomial-Dirichlet model centered at the *maximum a posteriori* (*map*) estimate of π. In brief, this is accomplished as follows: for each sample j, the marginal posterior distribution *p*(π_*j*_ | *Y*) is approximated as the posterior of a Bayesian multinomial Dirichlet model 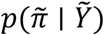 where 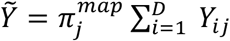 Here, the Dirichlet parameters α_*j*_ were all taken to be 0.5; this can be thought of as a probabilistic equivalent of using a pseudo-count of 0.5 yet also produced quantified uncertainty due to multivariate counting. The prior parameters were chosen as Θ = 0_([(*D*-1+*p*.]×*Q*)_, Γ = *I*_q_, and *v* = *D* + *P* + 9. Finally, we set the prior Ξ = (*v* - *D* - *P*) *BlockDiagonal* (*GG*^T^, *I*_*P*_.) where *G* is the (*D* - 1) × *D* ALR contrast matrix given by *G* = [*I*_*D-1*_ - 1]. This choice of Θ, Γ, Ξ, and *v* represents the weak prior belief that the correlation between the absolute abundance of taxa is, on average, small. This prior is closely related to the sparse penalization used by SparCC^61^. Using this model, priors, and inference, we sampled 2000 independent samples from the posterior distribution *p*(Λ, Σ|*Y, Z, X*).

Variable selection for the models was performed as described above in section (iii) and in Aho *et al*. (2019)^9^: for the three within-Parkinson’s covariance models (i.e. using only PD subjects at three taxonomic levels), the models were adjusted for COMT inhibitor medication use. For the three within-Controls covariance models we used intercept-only models. The matrix Σ represents a ([*D* - 1] + *Q*) × ([*D* - 1] + *Q*) covariance matrix encoding all possible covariances between ALR coordinates and metabolites. For model interpretation and inference, each posterior sample of the upper (*D* - 1) × *Q* submatrix was transformed to a *D* × *Q* matrix representing the covariance between microbial composition (now represented with respect to centered log-ratio coordinates) and metabolite abundances. Covariances were transformed to correlations using the function *cov2cor* in the R programming language. For the purposes of this study, we considered only those correlations that had a posterior mean equal to or larger than 0.3 and that had a 95% credible region not including zero. Conditioned on our chosen priors, this decision boundary can be thought of as limiting our results to correlations which we believe (with at least 95% certainty) are non-zero.

Correlation modelling was performed for bacterial genus, family, and phylum levels, using only metabolite abundance data at “Peak ID” level. This means that, although several of their corresponding MSI level 3 putative identifications were nominally the same, these were not merged before analysis, because they have different retention times and there is non-negligible uncertainty in their identification. After the correlations were calculated, we broadly assigned metabolite class information to these metabolites for Table 5 and Table 2 to aid in interpretation. These class assignments were then used to produce the Cytoscape^62^ network visualisations (v.3.8.0), both because classes simplify the networks and because they are more plausible than the putative MSI level 3 identifications. These classes were assigned by searching each putative identification of a metabolite feature against the Human Metabolome Database^63^ (HMDB) and the Kyoto Encyclopedia of Genes and Genomes^64^ (KEGG) entry.

## Data Availability

The 16S rRNA gene sequence abundance raw data is available from Aho et al. (2019)9. The metabolomics data will be hosted on MetaboLights (https://www.ebi.ac.uk/metabolights/index) and also on The University of Manchester servers. It will also be made available upon reasonable request. The clinical data is also available upon request due to European subject confidentiality laws.

## DATA AVAILABILITY

The 16S rRNA gene sequence abundance raw data is available from Aho *et al*. (2019)^9^. The metabolomics data will be hosted on MetaboLights (https://www.ebi.ac.uk/metabolights/index) and also on The University of Manchester servers. It will also be made available upon reasonable request. The clinical data is also available upon request due to European subject confidentiality laws.

## CODE AVAILABILITY

Code for the bacterial taxa-metabolite correlation analyses is available as R scripts’ files (supplementary files “Metabolomics.SERVER.SCRIPT.PD_ONLY.Selected.CORR.final” and “Metabolomics.SERVER.SCRIPT.CONTROLS_ONLY.Selected.CORR.final”). For metabolomics-only data analysis, deconvolution R scripts using XCMS and eRah, Matlab code for Partial Least Squares Discriminant Analysis (PLS-DA), R code for metabolite correlations, and Python code for Support Vector Machine (SVM) are available in the GitHub repository at github.com/drupadt/

## ACKNOWLEDGEMENTS

This study was funded by the Michael J. Fox Foundation for Parkinson’s Research, the Academy of Finland (295724, 310835), the Finnish Medical Foundation, and the Hospital District of Helsinki and Uusimaa (UAK1014004, UAK1014005, TYH2018224, TYH2020335).

DKT thanks Michael J Fox Foundation and Parkinson’s UK for supporting research leading to contribution to this work, and Prof Perdita Barran for her invaluable input throughout this project.

We thank Dr. Velma Aho for supporting this project by curating metadata from the Helsinki cohort, and for the R script for collapsing taxonomic levels.

## ETHICS DECLARATIONS

This study was conducted in accordance with the Declaration of Helsinki and was approved by the ethics committee of the Hospital District of Helsinki and Uusimaa. All participants gave informed consent.

## REFERENCES

1. Skjærbæk C, Knudsen K, Horsager J, Borghammer P. Gastrointestinal Dysfunction in Parkinson’s Disease. J Clin Med. 2021 Jan 31;10(3):493. doi: 10.3390/jcm10030493. PMID: 33572547; PMCID: PMC7866791.

2. Scheperjans F, Aho V, Pereira PA, Koskinen K, Paulin L, Pekkonen E, Haapaniemi E, Kaakkola S, Eerola-Rautio J, Pohja M, Kinnunen E, Murros K, Auvinen P. Gut microbiota are related to Parkinson’s disease and clinical phenotype. Mov Disord. 2015 Mar;30(3):350–8. doi: 10.1002/mds.26069. Epub 2014 Dec 5. PMID: 25476529.

3. Boertien JM, Pereira PAB, Aho VTE, Scheperjans F. Increasing Comparability and Utility of Gut Microbiome Studies in Parkinson’s Disease: A Systematic Review. J Parkinsons Dis. 2019;9(2):S297–S312. doi: 10.3233/JPD-191711. PMID: 31498131; PMCID: PMC6839453.

4. Cirstea MS, Yu AC, Golz E, Sundvick K, Kliger D, Radisavljevic N, Foulger LH, Mackenzie M, Huan T, Finlay BB, Appel-Cresswell S. Microbiota Composition and Metabolism Are Associated With Gut Function in Parkinson’s Disease. Mov Disord. 2020 Jul;35(7):1208–1217. doi: 10.1002/mds.28052. Epub 2020 May 1. PMID: 32357258.

5. Tan AH, Chong CW, Lim SY, Yap IKS, Teh CSJ, Loke MF, Song SL, Tan JY, Ang BH, Tan YQ, Kho MT, Bowman J, Mahadeva S, Yong HS, Lang AE. Gut Microbial Ecosystem in Parkinson Disease: New Clinicobiological Insights from Multi-Omics. Ann Neurol. 2021 Mar;89(3):546–559. doi: 10.1002/ana.25982. Epub 2021 Jan 11. PMID: 33274480.

6. Aho VTE, Houser MC, Pereira PAB, Chang J, Rudi K, Paulin L, Hertzberg V, Auvinen P, Tansey MG, Scheperjans F. Relationships of gut microbiota, short-chain fatty acids, inflammation, and the gut barrier in Parkinson’s disease. Mol Neurodegener. 2021 Feb 8;16(1):6. doi: 10.1186/s13024-021-00427-6. PMID: 33557896; PMCID: PMC7869249.

7. Unger MM, Spiegel J, Dillmann KU, Grundmann D, Philippeit H, Bürmann J, Faßbender K, Schwiertz A, Schäfer KH. Short chain fatty acids and gut microbiota differ between patients with Parkinson’s disease and age-matched controls. Parkinsonism Relat Disord. 2016 Nov;32:66–72. doi: 10.1016/j.parkreldis.2016.08.019. Epub 2016 Aug 26. PMID: 27591074.

8. Mertsalmi TH, Aho VTE, Pereira PAB, Paulin L, Pekkonen E, Auvinen P, Scheperjans F. More than constipation - bowel symptoms in Parkinson’s disease and their connection to gut microbiota. Eur J Neurol. 2017 Nov;24(11):1375–1383. doi: 10.1111/ene.13398. Epub 2017 Sep 11. PMID: 28891262.

9. Aho VTE, Pereira PAB, Voutilainen S, Paulin L, Pekkonen E, Auvinen P, Scheperjans F. Gut microbiota in Parkinson’s disease: Temporal stability and relations to disease progression. EBioMedicine. 2019 Jun;44:691–707. doi: 10.1016/j.ebiom.2019.05.064. Epub 2019 Jun 18. PMID: 31221587; PMCID: PMC6606744.

10. Pereira PAB, Aho VTE, Paulin L, Pekkonen E, Auvinen P, Scheperjans F. Oral and nasal microbiota in Parkinson’s disease. Parkinsonism Relat Disord. 2017 May;38:61–67. doi: 10.1016/j.parkreldis.2017.02.026. Epub 2017 Feb 22. PMID: 28259623.

11. Wishart DS, Feunang YD, Marcu A, Guo AC, Liang K, Vázquez-Fresno R, Sajed T, Johnson D, Li C, Karu N, Sayeeda Z, Lo E, Assempour N, Berjanskii M, Singhal S, Arndt D, Liang Y, Badran H, Grant J, Serra-Cayuela A, Liu Y, Mandal R, Neveu V, Pon A, Knox C, Wilson M, Manach C, Scalbert A. HMDB 4.0: the human metabolome database for 2018. Nucleic Acids Res. 2018 Jan 4;46(D1):D608–D617. doi: 10.1093/nar/gkx1089. PMID: 29140435; PMCID: PMC5753273.

12. Sud M, Fahy E, Cotter D, Brown A, Dennis EA, Glass CK, Merrill AH Jr, Murphy RC, Raetz CR, Russell DW, Subramaniam S. LMSD: LIPID MAPS structure database. Nucleic Acids Res. 2007 Jan;35(Database issue):D527–32. doi: 10.1093/nar/gkl838. Epub 2006 Nov 10. PMID: 17098933; PMCID: PMC1669719.

13. Sumner LW, Amberg A, Barrett D, Beale MH, Beger R, Daykin CA, Fan TW, Fiehn O, Goodacre R, Griffin JL, Hankemeier T, Hardy N, Harnly J, Higashi R, Kopka J, Lane AN, Lindon JC, Marriott P, Nicholls AW, Reily MD, Thaden JJ, Viant MR. Proposed minimum reporting standards for chemical analysis Chemical Analysis Working Group (CAWG) Metabolomics Standards Initiative (MSI). Metabolomics. 2007 Sep;3(3):211–221. doi: 10.1007/s11306-007-0082-2. PMID: 24039616; PMCID: PMC3772505.

14. Sinclair E, Trivedi DK, Sarkar D, Walton-Doyle C, Milne J, Kunath T, Rijs AM, de Bie RMA, Goodacre R, Silverdale M, Barran P. Metabolomics of sebum reveals lipid dysregulation in Parkinson’s disease. Nat Commun. 2021 Mar 11;12(1):1592. doi: 10.1038/s41467-021-21669-4. PMID: 33707447; PMCID: PMC7952564.

15. Tan AH, Chong CW, Lim SY, Yap IKS, Teh CSJ, Loke MF, Song SL, Tan JY, Ang BH, Tan YQ, Kho MT, Bowman J, Mahadeva S, Yong HS, Lang AE. Gut Microbial Ecosystem in Parkinson Disease: New Clinicobiological Insights from Multi-Omics. Ann Neurol. 2021 Mar;89(3):546–559. doi: 10.1002/ana.25982. Epub 2021 Jan 11. PMID: 33274480.

16. Rattray NJW, Trivedi DK, Xu Y, Chandola T, Johnson CH, Marshall AD, Mekli K, Rattray Z, Tampubolon G, Vanhoutte B, White IR, Wu FCW, Pendleton N, Nazroo J, Goodacre R. Metabolic dysregulation in vitamin E and carnitine shuttle energy mechanisms associate with human frailty. Nat Commun. 2019 Nov 5;10(1):5027. doi: 10.1038/s41467-019-12716-2. PMID: 31690722; PMCID: PMC6831565.

17. Alecu I, Bennett SAL. Dysregulated Lipid Metabolism and Its Role in α-Synucleinopathy in Parkinson’s Disease. Front Neurosci. 2019 Apr 11;13:328. doi: 10.3389/fnins.2019.00328. PMID: 31031582; PMCID: PMC6470291.

18. Lin G, Wang L, Marcogliese PC, Bellen HJ. Sphingolipids in the Pathogenesis of Parkinson’s Disease and Parkinsonism. Trends Endocrinol Metab. 2019 Feb;30(2):106–117. doi: 10.1016/j.tem.2018.11.003. Epub 2018 Dec 6. PMID: 30528460.

19. Hallett PJ, Engelender S, Isacson O. Lipid and immune abnormalities causing age-dependent neurodegeneration and Parkinson’s disease. J Neuroinflammation. 2019 Jul 22;16(1):153. doi: 10.1186/s12974-019-1532-2. PMID: 31331333; PMCID: PMC6647317.

20. Xicoy H, Wieringa B, Martens GJM. The Role of Lipids in Parkinson’s Disease. Cells. 2019 Jan 7;8(1):27. doi: 10.3390/cells8010027. PMID: 30621069; PMCID: PMC6356353.

21. Saiki S, Hatano T, Fujimaki M, Ishikawa KI, Mori A, Oji Y, Okuzumi A, Fukuhara T, Koinuma T, Imamichi Y, Nagumo M, Furuya N, Nojiri S, Amo T, Yamashiro K, Hattori N. Decreased long-chain acylcarnitines from insufficient β-oxidation as potential early diagnostic markers for Parkinson’s disease. Sci Rep. 2017 Aug 4;7(1):7328. doi: 10.1038/s41598-017-06767-y. PMID: 28779141; PMCID: PMC5544708.

22. Rosario D, Bidkhori G, Lee S, Bedarf J, Hildebrand F, Le Chatelier E, Uhlen M, Ehrlich SD, Proctor G, Wüllner U, Mardinoglu A, Shoaie S. Systematic analysis of gut microbiome reveals the role of bacterial folate and homocysteine metabolism in Parkinson’s disease. Cell Rep. 2021 Mar 2;34(9):108807. doi: 10.1016/j.celrep.2021.108807. PMID: 33657381.

23. da Silva RP, Kelly KB, Al Rajabi A, Jacobs RL. Novel insights on interactions between folate and lipid metabolism. Biofactors. 2014 May-Jun;40(3):277–83. doi: 10.1002/biof.1154. Epub 2013 Dec 18. PMID: 24353111; PMCID: PMC4153959.

24. Shao Y, Li T, Liu Z, Wang X, Xu X, Li S, Xu G, Le W. Comprehensive metabolic profiling of Parkinson’s disease by liquid chromatography-mass spectrometry. Mol Neurodegener. 2021 Jan 23;16(1):4. doi: 10.1186/s13024-021-00425-8. PMID: 33485385; PMCID: PMC7825156.

25. Zhao, H.; Wang, C.; Zhao, N.; Li, W.; Yang, Z.; Liu, X.; Le, W.; Zhang, X. Potential biomarkers of Parkinson’s disease revealed by plasma metabolic profiling. J. Chromatogr. B Anal. Technol. Biomed. Life Sci. 2018, 1081–1082, 101–108

26. Crooks SA, Bech S, Halling J, Christiansen DH, Ritz B, Petersen MS. Carnitine levels and mutations in the SLC22A5 gene in Faroes patients with Parkinson’s disease. Neurosci Lett. 2018 May 14;675:116–119. doi: 10.1016/j.neulet.2018.03.064. Epub 2018 Mar 31. PMID: 29614331.

27. Jiménez-Jiménez FJ, Rubio JC, Molina JA, Martín MA, Campos Y, Benito-León J, Ortí-Pareja M, Gasalla T, Arenas J. Cerebrospinal fluid carnitine levels in patients with Parkinson’s disease. J Neurol Sci. 1997 Feb 12;145(2):183–5. doi: 10.1016/s0022-510x(96)00259-6. PMID: 9094047.

28. Lin G, Lee PT, Chen K, Mao D, Tan KL, Zuo Z, Lin WW, Wang L, Bellen HJ. Phospholipase PLA2G6, a Parkinsonism-Associated Gene, Affects Vps26 and Vps35, Retromer Function, and Ceramide Levels, Similar to α-Synuclein Gain. Cell Metab. 2018 Oct 2;28(4):605–618.e6. doi: 10.1016/j.cmet.2018.05.019. Epub 2018 Jun 14. PMID: 29909971.

29. Belarbi K, Cuvelier E, Bonte MA, Desplanque M, Gressier B, Devos D, Chartier-Harlin MC. Glycosphingolipids and neuroinflammation in Parkinson’s disease. Mol Neurodegener. 2020 Oct 17;15(1):59. doi: 10.1186/s13024-020-00408-1. PMID: 33069254; PMCID: PMC7568394.

30. Zhu M, Li J, Fink AL. The association of alpha-synuclein with membranes affects bilayer structure, stability, and fibril formation. J Biol Chem. 2003 Oct 10;278(41):40186–97. doi: 10.1074/jbc.M305326200. Epub 2003 Jul 28. PMID: 12885775.

31. Madine J, Doig AJ, Middleton DA. A study of the regional effects of alpha-synuclein on the organization and stability of phospholipid bilayers. Biochemistry. 2006 May 9;45(18):5783–92. doi: 10.1021/bi052151q. PMID: 16669622.

32. Rawat A, Langen R, Varkey J. Membranes as modulators of amyloid protein misfolding and target of toxicity. Biochim Biophys Acta Biomembr. 2018 Sep;1860(9):1863–1875. doi: 10.1016/j.bbamem.2018.04.011. Epub 2018 Apr 25. PMID: 29702073; PMCID: PMC6203680.

33. Jo E, McLaurin J, Yip CM, St George-Hyslop P, Fraser PE. alpha-Synuclein membrane interactions and lipid specificity. J Biol Chem. 2000 Nov 3;275(44):34328–34. doi: 10.1074/jbc.M004345200. PMID: 10915790.

34. Broersen K, van den Brink D, Fraser G, Goedert M, Davletov B. Alpha-synuclein adopts an alpha-helical conformation in the presence of polyunsaturated fatty acids to hinder micelle formation. Biochemistry. 2006 Dec 26;45(51):15610–6. doi: 10.1021/bi061743l. Epub 2006 Dec 7. PMID: 17176082.

35. De Franceschi G, Frare E, Bubacco L, Mammi S, Fontana A, de Laureto PP. Molecular insights into the interaction between alpha-synuclein and docosahexaenoic acid. J Mol Biol. 2009 Nov 20;394(1):94–107. doi: 10.1016/j.jmb.2009.09.008. Epub 2009 Sep 8. PMID: 19747490.

36. De Franceschi G, Frare E, Pivato M, Relini A, Penco A, Greggio E, Bubacco L, Fontana A, de Laureto PP. Structural and morphological characterization of aggregated species of α-synuclein induced by docosahexaenoic acid. J Biol Chem. 2011 Jun 24;286(25):22262–74. doi: 10.1074/jbc.M110.202937. Epub 2011 Apr 28. PMID: 21527634; PMCID: PMC3121372.

37. Ludtmann MH, Angelova PR, Ninkina NN, Gandhi S, Buchman VL, Abramov AY. Monomeric Alpha-Synuclein Exerts a Physiological Role on Brain ATP Synthase. J Neurosci. 2016 Oct 12;36(41):10510–10521. doi: 10.1523/JNEUROSCI.1659-16.2016. PMID: 27733604; PMCID: PMC5059426.

38. Henzi V, Reichling DB, Helm SW, MacDermott AB. L-proline activates glutamate and glycine receptors in cultured rat dorsal horn neurons. Mol Pharmacol. 1992 Apr;41(4):793–801. PMID: 1349155.

39. Wu G, Bazer FW, Datta S, Johnson GA, Li P, Satterfield MC, Spencer TE. Proline metabolism in the conceptus: implications for fetal growth and development. Amino Acids. 2008 Nov;35(4):691–702. doi: 10.1007/s00726-008-0052-7. Epub 2008 Mar 11. PMID: 18330497.

40. Ji Y, Guo Q, Yin Y, Blachier F, Kong X. Dietary proline supplementation alters colonic luminal microbiota and bacterial metabolite composition between days 45 and 70 of pregnancy in Huanjiang mini-pigs. J Anim Sci Biotechnol. 2018 Jan 30;9:18. doi: 10.1186/s40104-018-0233-5. PMID: 29423216; PMCID: PMC5789534.

41. Dunn WB, Broadhurst D, Begley P, Zelena E, Francis-McIntyre S, Anderson N, Brown M, Knowles JD, Halsall A, Haselden JN, Nicholls AW, Wilson ID, Kell DB, Goodacre R; Human Serum Metabolome (HUSERMET) Consortium. Procedures for large-scale metabolic profiling of serum and plasma using gas chromatography and liquid chromatography coupled to mass spectrometry. Nat Protoc. 2011 Jun 30;6(7):1060–83. doi: 10.1038/nprot.2011.335. PMID: 21720319.

42. Begley P, Francis-McIntyre S, Dunn WB, Broadhurst DI, Halsall A, Tseng A, Knowles J; HUSERMET Consortium,, Goodacre R, Kell DB. Development and performance of a gas chromatography-time-of-flight mass spectrometry analysis for large-scale nontargeted metabolomic studies of human serum. Anal Chem. 2009 Aug 15;81(16):7038–46. doi: 10.1021/ac9011599. PMID: 19606840.

43. Chambers MC, Maclean B, Burke R, Amodei D, Ruderman DL, Neumann S, Gatto L, Fischer B, Pratt B, Egertson J, Hoff K, Kessner D, Tasman N, Shulman N, Frewen B, Baker TA, Brusniak MY, Paulse C, Creasy D, Flashner L, Kani K, Moulding C, Seymour SL, Nuwaysir LM, Lefebvre B, Kuhlmann F, Roark J, Rainer P, Detlev S, Hemenway T, Huhmer A, Langridge J, Connolly B, Chadick T, Holly K, Eckels J, Deutsch EW, Moritz RL, Katz JE, Agus DB, MacCoss M, Tabb DL, Mallick P. A cross-platform toolkit for mass spectrometry and proteomics. Nat Biotechnol. 2012 Oct;30(10):918–20. doi: 10.1038/nbt.2377. PMID: 23051804; PMCID: PMC3471674.

44. RStudio Team. RStudio: Integrated Development Environment for R. Boston, MA; 2015. Available from: http://www.rstudio.com/

45. Smith CA, Want EJ, O’Maille G, Abagyan R, Siuzdak G. XCMS: processing mass spectrometry data for metabolite profiling using nonlinear peak alignment, matching, and identification. Anal Chem. 2006 Feb 1;78(3):779–87. doi: 10.1021/ac051437y. PMID: 16448051.

46. Xavier Domingo-Almenara, Jesus Brezmes, Maria Vinaixa, Sara Samino, Noelia Ramirez, Marta Ramon-Krauel, Carles Lerin, Marta Díaz, Lourdes Ibáñez, Xavier Correig, Alexandre Perera-Lluna, and Oscar Yanes. eRah: A Computational Tool Integrating Spectral Deconvolution and Alignment with Quantification and Identification of Metabolites in GC/MS-Based Metabolomics. Analytical Chemistry. 2016 88 (19), 9821–9829. DOI: 10.1021/acs.analchem.6b02927

47. Duan KB, Rajapakse JC, Wang H, Azuaje F. Multiple SVM-RFE for gene selection in cancer classification with expression data. IEEE Trans Nanobioscience. 2005 Sep;4(3):228–34. doi: 10.1109/tnb.2005.853657. PMID: 16220686.

48. Kurlowicz, L., & Greenberg, S. A. (2007). The geriatric depression scale (GDS). AJN The American Journal of Nursing, 107(10), 67–68.

49. Ombaugh, T. N., McDowell, I., Kristjansson, B., & Hubley, A. M. (1996). Mini-Mental State Examination (MMSE) and the Modified MMSE (3MS): A psychometric comparison and normative data. Psychological Assessment, 8(1), 48–59.

50. Chaudhuri KR, Martinez-Martin P. Quantitation of non-motor symptoms in Parkinson’s disease. Eur J Neurol. 2008 Sep;15 Suppl 2:2–7. doi: 10.1111/j.1468-1331.2008.02212.x. PMID: 18702736.

51. Stiasny-Kolster K, Mayer G, Schäfer S, Möller JC, Heinzel-Gutenbrunner M, Oertel WH. The REM sleep behavior disorder screening questionnaire--a new diagnostic instrument. Mov Disord. 2007 Dec;22(16):2386–93. doi: 10.1002/mds.21740. PMID: 17894337.

52. Rome Foundation. Guidelines--Rome III Diagnostic Criteria for Functional Gastrointestinal Disorders. J Gastrointestin Liver Dis. 2006 Sep;15(3):307–12. PMID: 17203570.

53. Agachan F, Chen T, Pfeifer J, Reissman P, Wexner SD. A constipation scoring system to simplify evaluation and management of constipated patients. Dis Colon Rectum. 1996 Jun;39(6):681–5. doi: 10.1007/BF02056950. PMID: 8646957.

54. Perez Lloret S, Pirán Arce G, Rossi M, Caivano Nemet ML, Salsamendi P, Merello M. Validation of a new scale for the evaluation of sialorrhea in patients with Parkinson’s disease. Mov Disord. 2007 Jan;22(1):107–11. doi: 10.1002/mds.21152. PMID: 17089393.

55. Lam K, Lam FK, Lau KK, et al. Simple clinical tests may predict severe oropharyngeal dysphagia in Parkinson’s disease. Movement Disorders : Official Journal of the Movement Disorder Society. 2007 Apr;22(5):640–644. DOI: 10.1002/mds.21362.

56. Li S, Park Y, Duraisingham S, Strobel FH, Khan N, Soltow QA, Jones DP, Pulendran B. Predicting network activity from high throughput metabolomics. PLoS Comput Biol. 2013;9(7):e1003123. doi: 10.1371/journal.pcbi.1003123. Epub 2013 Jul 4. PMID: 23861661; PMCID: PMC3701697.

57. Chong J, Xia J. MetaboAnalystR: an R package for flexible and reproducible analysis of metabolomics data. Bioinformatics. 2018 Dec 15;34(24):4313–4314. doi: 10.1093/bioinformatics/bty528. PMID: 29955821; PMCID: PMC6289126.

58. Silverman, JD, Roche, K, Holmes, ZC, David, LA, and Mukherjee, S. Bayesian Multinomial Logistic Normal Models through Marginally Latent Matrix-T Processes. 2019, arXiv e-prints, 1903.11695

59. R Core Team (2020). R: A language and environment for statistical computing. R Foundation for Statistical Computing, Vienna, Austria. URL https://www.R-project.org/.

60. Aitchison, J. (1986), The statistical analysis of compositional data, Monographs on statistics and applied probability, Chapman and Hall, London; New York.

61. Friedman J, Alm EJ. Inferring correlation networks from genomic survey data. PLoS Comput Biol. 2012;8(9):e1002687. doi: 10.1371/journal.pcbi.1002687. Epub 2012 Sep 20. PMID: 23028285; PMCID: PMC3447976.

62. Shannon P, Markiel A, Ozier O, Baliga NS, Wang JT, Ramage D, Amin N, Schwikowski B, Ideker T. Cytoscape: a software environment for integrated models of biomolecular interaction networks. Genome Res. 2003 Nov;13(11):2498–504. doi: 10.1101/gr.1239303. PMID: 14597658; PMCID: PMC403769.

63. Wishart DS, Tzur D, Knox C, Eisner R, Guo AC, Young N, Cheng D, Jewell K, Arndt D, Sawhney S, Fung C, Nikolai L, Lewis M, Coutouly MA, Forsythe I, Tang P, Shrivastava S, Jeroncic K, Stothard P, Amegbey G, Block D, Hau DD, Wagner J, Miniaci J, Clements M, Gebremedhin M, Guo N, Zhang Y, Duggan GE, Macinnis GD, Weljie AM, Dowlatabadi R, Bamforth F, Clive D, Greiner R, Li L, Marrie T, Sykes BD, Vogel HJ, Querengesser L. HMDB: the Human Metabolome Database. Nucleic Acids Res. 2007 Jan;35(Database issue):D521–6. doi: 10.1093/nar/gkl923. PMID: 17202168; PMCID: PMC1899095.

64. Kanehisa M, Goto S. KEGG: kyoto encyclopedia of genes and genomes. Nucleic Acids Res. 2000 Jan 1;28(1):27–30. doi: 10.1093/nar/28.1.27. PMID: 10592173; PMCID: PMC102409.

