## Supplementary material for "Multiomics implicate gut microbiota in altered lipid and energy metabolism in Parkinson’s disease": Metabolomics.SERVER.SCRIPT.CONTROLS_ONLY.Selected.CORR.final.R

```
library(dplyr)
library(phyloseq)
# devtools::install_github("jsilve24/stray")
library(stray)
# devtools::install_github("jsilve24/driver")
library(driver)
source("taxaLevelCollapser_v5.R")


otu <- read.csv("pdfu_16s_OTU_counts_METABOLOMICS.2.csv", sep = ";", header = TRUE,
                row.names = 1, strip.white = TRUE)

tax <- read.csv("pdfu_16s_taxonomy.csv", sep = ",", header=TRUE,
                row.names = 1, strip.white = TRUE)

sampledata <- read.csv("Metabolomics_metadata_ready.4.csv", sep = ",", dec = ".", 
                       header = TRUE, row.names = 1,
                       strip.white = TRUE)


sampledata.2 <- sampledata


otu.2 <- t(otu)

otu.matrix <- as.matrix(otu.2)
tax.matrix <- as.matrix(tax)

dim(otu.2)
dim(tax)


otu.table <- otu_table(otu.matrix, taxa_are_rows = TRUE)
tax.table <- tax_table(tax.matrix)
sampledata.3 <- sample_data(sampledata.2)

dim(sampledata.3)


# 124 samples total:
physeqfinal <- phyloseq(otu.table, tax.table, sampledata.3)
physeqfinal

sample_data(physeqfinal)
str(sample_data(physeqfinal))
str(sampledata.2)


### Get only the CONTROL samples:
table(sample_data(physeqfinal)$Group)
physeqfinal <- subset_samples(physeqfinal, Group == "C")
table(sample_data(physeqfinal)$Group)
sample_data(physeqfinal)


######### METABOLITES:

# We are only using those metabolites that were diferentially abundant between C and PD (i.e. "selected").
# We randomize the order of the metabolites (i.e. columns) to make sure that no biases are introduced by
# using the metabolites in the original order of the table (they have different mass spectrometry sources).
# We will save that object (to be on the safe side):

#set.seed(666)
#head(GC)
#GC.2 <- GC[ ,sample(ncol(GC))]
#head(GC.2)
#write.table(GC.2, file = "GC.rand.selected.tsv", col.names = NA)
# Then separate the object into PD or Controls, as desired. I already had those files prepared,
# so I don't do that here; I just load up a ready-made file. Files are available upon request, including RAW.


GC.2 <- read.csv("GC.rand.selected.CONTROLS_ONLY.csv", sep = "", dec = ".", header = TRUE,
               row.names = 1, strip.white = TRUE)


#### Log 10 Transform:
any(GC.2 == 0)
GC.log <- log10(1 + GC.2)
GC.log


#### Z-transform to put all combined MS datasets in the same scale:
GC.log.Z <- as.data.frame(apply(GC.log, 2, scale))
GC.log.Z
colMeans(GC.log.Z)
sapply(GC.log.Z, sd)

rownames(GC.log.Z)
rownames(GC.log.Z) <- rownames(GC.2)

GC.3 <- t(GC.log.Z)
GC.4 <- as.matrix(GC.3)
rownames(GC.4)
GC.4


######### Back to TAXA.
# The script for collapsing taxonomic levels was writen by Dr Velma Aho.
# After collapsing the taxonomic levels, amalgamate those TAXA
# that don’t pass filtering to a category called “other”
# to maintain the proper variance in the multinomial model.
# Choose the right option for the desired taxonomic level:

physeqfinal.2 <-collapseTaxLevel(physeqfinal, level="Genus")
physeqfinal.2
physeqfinal
physeqfinal.3 <- physeqfinal.2

keep_ix <- taxa_sums(physeqfinal.3) > 4
keep_ix <- keep_ix & (rowSums(otu_table(physeqfinal.3)>2)>3)
physeqfinal.3 <- merge_taxa(physeqfinal.3, taxa_names(physeqfinal.3)[!keep_ix])
nms <- taxa_names(physeqfinal.3)
rnm <- which(taxa_names(physeqfinal.3) == taxa_names(physeqfinal.3)[!keep_ix][1])
nms[rnm] <- "other"
taxa_names(physeqfinal.3) <- nms
rm(nms, rnm)

physeqfinal
physeqfinal.2
physeqfinal.3


Y <- otu_table(physeqfinal.3, taxa_are_rows = TRUE)

Z <- GC.4

sample_dat <- as.data.frame(as(sample_data(physeqfinal.3), "matrix"))


# We use a model with intercept only:
X.check.2 <- model.matrix(~ 1, data = sample_dat)
X.check.2

X <- t(model.matrix(~ 1, data = sampledata.ref))

# save dimensions for easy reference:
N <- ncol(Y) 
D <- nrow(Y)
P <- nrow(Z)
Q <- nrow(X)

dim(Y)
dim(Z)
dim(X)


# Priors (see our paper for an explanation of what we are doing here and why):

upsilon <- (D-1+P)+10
Xi <- diag(D-1+P)
GG <- cbind(diag(D-1), -1)
Xi[1:(D-1), 1:(D-1)] <- GG %*% diag(D) %*% t(GG)
Xi <- Xi * (upsilon-D-P)

Gamma <- diag(Q)

Theta <- matrix(0, D-1+P, 1)

dim(Gamma)
dim(Theta)


### Fitting the model:

set.seed(666)


fit <- orthus(Y, Z, X, Theta = Theta, Gamma = Gamma, Xi = Xi, upsilon = upsilon,
              calcGradHess = FALSE,
              multDirichletBoot = .5,
              n_samples = 2000,
              verbose = TRUE)


fit.2 <- to_clr(fit)
#print(fit.2)


# Now get those correlations from the covariances:

xcor.2 <- fit.2$Sigma
xcor.2
is.array(xcor.2)
dim(xcor.2)


for (i in 1:dim(xcor.2)[3]){
  xcor.2[,,i] <- cov2cor(xcor.2[,,i])
}


xcor.3 <- xcor.2[1:D,(D+1):(D+P),]
xcor.3
dim(xcor.3)


xcor.summary.COR <- gather_array(xcor.3, cov, taxa, metabolite, iter) %>%
  mutate(tm = paste0(taxa, "_", metabolite)) %>% 
  mutate(taxa = rownames(Y)[taxa], metabolite = rownames(Z)[metabolite]) %>% 
  group_by(taxa, metabolite) %>% 
  summarise_posterior(cov) %>% 
  arrange(mean) %>% 
  filter(taxa != 'other')

res.cor <- xcor.summary.COR %>% 
  filter(sign(p2.5) == sign(p97.5)) %>% 
  filter(abs(mean) > 0.3) # or whatever correlation we want as a minimum

res.cor

write.table(res.cor, file = "res.coR.selected.CONTROLS_ONLY.GENUS.tsv", col.names = NA)


sessionInfo()

# R version 3.6.0 (2019-04-26)
# Platform: x86_64-apple-darwin15.6.0 (64-bit)
# Running under: macOS  10.16
# 
# Matrix products: default
# LAPACK: /Library/Frameworks/R.framework/Versions/3.6/Resources/lib/libRlapack.dylib
# 
# locale:
#   [1] en_US.UTF-8/en_US.UTF-8/en_US.UTF-8/C/en_US.UTF-8/en_US.UTF-8
# 
# attached base packages:
#   [1] stats     graphics  grDevices utils     datasets  methods   base     
# 
# other attached packages:
#   [1] driver_0.1.1    stray_0.1.13    phyloseq_1.27.6 dplyr_0.8.4    
# 
# loaded via a namespace (and not attached):
#   [1] tidyselect_1.0.0    purrr_0.3.3         reshape2_1.4.3     
# [4] splines_3.6.0       lattice_0.20-38     rhdf5_2.27.19      
# [7] colorspace_1.4-1    vctrs_0.3.1         stats4_3.6.0       
# [10] mgcv_1.8-28         survival_2.44-1.1   rlang_0.4.10       
# [13] pillar_1.4.3        glue_1.3.1          BiocGenerics_0.29.2
# [16] foreach_1.4.4       lifecycle_0.2.0     plyr_1.8.5         
# [19] stringr_1.4.0       zlibbioc_1.29.0     Biostrings_2.51.5  
# [22] munsell_0.5.0       gtable_0.3.0        coda_0.19-3        
# [25] codetools_0.2-16    forcats_0.4.0       Biobase_2.43.1     
# [28] permute_0.9-5       IRanges_2.17.5      biomformat_1.11.1  
# [31] parallel_3.6.0      Rcpp_1.0.3          arrayhelpers_1.1-0 
# [34] scales_1.1.0        vegan_2.5-4         S4Vectors_0.21.24  
# [37] jsonlite_1.6        XVector_0.23.2      tidybayes_2.0.1    
# [40] svUnit_0.7-12       ggplot2_3.2.1       stringi_1.4.5      
# [43] grid_3.6.0          ade4_1.7-13         tools_3.6.0        
# [46] magrittr_1.5        lazyeval_0.2.2      tibble_3.0.1       
# [49] cluster_2.0.8       tidyr_1.0.2         crayon_1.3.4       
# [52] ape_5.3             pkgconfig_2.0.3     MASS_7.3-51.4      
# [55] ellipsis_0.3.0      Matrix_1.2-17       data.table_1.12.6  
# [58] assertthat_0.2.1    rstudioapi_0.10     iterators_1.0.10   
# [61] Rhdf5lib_1.5.4      R6_2.4.1            multtest_2.39.0    
# [64] igraph_1.2.4.1      nlme_3.1-139        compiler_3.6.0  
```
