## Supplementary material for "Multiomics implicate gut microbiota in altered lipid and energy metabolism in Parkinson’s disease": taxaLevelCollapser_v5.R

```
##########################################################################
## Function for collapsing phyloseq objects to lower taxonomical levels ##
##########################################################################

## VTEA, 14 December 2018

# Changes from previous versions:
# v3 : fixed script to work even if taxa_are_rows = TRUE
# v4 : changed the behavior of the fixUnclassifieds argument, see below.
# v5 : overall streamlined code; switched from for-loops to apply-family alternatives

# Dependencies: packages phyloseq and reshape2

# Required input:
# Phyloseq-object and the name of a taxonomical level
# eg. collapseTaxLevel(phylo_object, "Family")

# Otuput:
# A new phyloseq object that represents the selected level.

# Unclassified taxa:
# By default, renames unclassified taxa to indicate the lowest unclassified level, 
# e.g. "Actinobacteria_unclassified_Order" etc.
# If you would rather collapse all the unclassified taxa into one bin on the target level, 
# which looks cleaner, but loses some taxonomy information,
# use fixUnclassifieds = FALSE.

collapseTaxLevel <- function(phylo_obj, level, fixUnclassifieds = TRUE, maxUnclassifiedLevel = NA){
	
	library(reshape2)
	library(phyloseq)
	
	unclassifiedFix <- function(phylo_obj){
	  taxmat <- as(tax_table(phylo_obj), "matrix")
	  taxmat <- t(apply(taxmat, 1,
	                    function(x)
	                      if(length(grep("unclassified", x)) > 0) {
	                        x <- gsub("unclassified",
	                                  paste("(unclassified", "_",
	                                        tolower(names(x)[min(grep("unclassified", x, ignore.case = TRUE))]),
	                                        ")", sep = ""), x)
	                      } else {x <- x}
	  ))
	  phylo_obj <- makeNewPhyloObj(phylo_obj, otu_table(phylo_obj), taxmat)
	  return(phylo_obj)		
	}
	
	collapseOtuTable <- function(phylo_obj, level){
		tax <- as.data.frame(as(tax_table(phylo_obj), "matrix"))
		otu <- as.data.frame(as(otu_table(phylo_obj), "matrix"))
		otu[, level] <- tax[, level]
		otu_collapsed <- melt(otu, id = (level))
		otu_collapsed <- acast(otu_collapsed, as.formula(paste(level, "~variable", sep = "")),  sum)
		otu_collapsed <- otu_collapsed[order(rownames(otu_collapsed)), ]
		return(otu_collapsed)
	}
	
	makeNewPhyloObj <- function(phylo_obj, otutable,  taxtable){
		if (length(phylo_obj@sam_data) == 0) {
			newPhyloObj <- phyloseq(otu_table(otutable,  taxa_are_rows = TRUE), tax_table(taxtable))		
		} else {	
			newPhyloObj <- phyloseq(otu_table(otutable,  taxa_are_rows = TRUE), tax_table(taxtable), sample_data(phylo_obj))
		}
		return(newPhyloObj)	
	}
	
	# the entire script was written assuming taxa_are_rows == TRUE -> fix
	if (taxa_are_rows(phylo_obj) == FALSE) {
	
			phylo_obj <- phyloseq(tax_table(phylo_obj),
			                      sample_data(phylo_obj),
			                      otu_table(t(otu_table(phylo_obj))), taxa_are_rows = TRUE)
	}
		
	levelNum <- grep(level, colnames(tax_table(phylo_obj)))
	
	if (length(levelNum) == 0){
		
		stop("Incorrect level! Use taxonomical levels that are present in your data.")
		
	} else if (nlevels(factor(tax_table(phylo_obj)[, level])) == 1) {
		
		stop("Only one taxon on this level, nothing to collapse!")
		
	} else if (levelNum == 1){
		
		otu_collapsed <- collapseOtuTable(phylo_obj, level)
		
		tax_collapsed <- data.frame(rownames(otu_collapsed), row.names = rownames(otu_collapsed))
		colnames(tax_collapsed) <- level
		
	} else {
	  
	  if (fixUnclassifieds == FALSE) {
		tax <- as(tax_table(phylo_obj), "matrix")
		tax <- apply(tax, 2, function(x) gsub(".*_unclassified", "unclassified", x))
		phylo_obj <- phyloseq(otu_table(phylo_obj), sample_data(phylo_obj), tax_table(tax))
	  }
	  
	  else {
	    phylo_obj <- unclassifiedFix(phylo_obj)
	  }

		otu_collapsed <- collapseOtuTable(phylo_obj, level)
		
		tax <- as(tax_table(phylo_obj), "matrix")
		tax_collapsed <- unique(tax[, 1:levelNum])
		
		if (fixUnclassifieds == FALSE) {
		tax_collapsed <- tax_collapsed[-grep("unclassified", tax_collapsed[,levelNum]),]
		tax_collapsed <- rbind(tax_collapsed, rep("unclassified", (levelNum)))
		}
		
		rownames(tax_collapsed) <- tax_collapsed[, level]
		tax_collapsed <- tax_collapsed[order(rownames(tax_collapsed)), ]
		
		if(!is.na(maxUnclassifiedLevel)){
			
			ucAdd <- sort(unique(unlist(sapply(colnames(tax_collapsed)[1:(which(colnames(tax_collapsed) == maxUnclassifiedLevel) - 1)],
			                 function (x) grep("unclassified", tax_collapsed[, x])))))
			
			if(length(ucAdd) == 1){
				ucCounts <- otu_collapsed[ucAdd, ]
			} else {
				ucCounts <- colSums(otu_collapsed[ucAdd, ])
			}
			
			if(length(grep("^unclassified$", rownames(otu_collapsed)) > 0)){
				ucCounts <- ucCounts + otu_collapsed["unclassified", ]
				otu_collapsed <- otu_collapsed[-ucAdd, ]
				otu_collapsed["unclassified", ] <- ucCounts
				tax_collapsed <- tax_collapsed[-ucAdd, ]
			} else {
				otu_collapsed <- rbind(otu_collapsed[-ucAdd, ], unclassified = ucCounts)
				tax_collapsed <- rbind(tax_collapsed[-ucAdd, ], unclassified = rep("unclassified", length = ncol(tax_collapsed)))
			}
		}
	}
	
	new_phylo_obj <- makeNewPhyloObj(phylo_obj, otu_collapsed, tax_collapsed)
	
	return(new_phylo_obj)
	
}
```
